# Evaluation of a Contactless Sleep Monitoring Device for Sleep Stage Detection against Home Polysomnography in a Healthy Population

**DOI:** 10.1101/2025.05.06.25326860

**Authors:** Marie-Ange Stefanos, Guillaume De Laboulaye, David Campo, Martin De Gourcuff, Pierre Escourrou, Boris Matrot, Anna Sigridur Islind, Pierre A. Geoffroy

## Abstract

**Background:** Sleep is essential for overall health and well-being, but assessing sleep architecture and quality is often costly and time-consuming, relying primarily on polysomnography (PSG). Wearable and nearable devices offer potential alternatives, but they regularly lack rigorous validation, especially in real-world settings.

**Objectives:** This study evaluates the accuracy and reliability of Withings Sleep Analyzer (WSA), a contactless sleep monitoring device, compared to PSG in a home setting using a large and diverse cohort of healthy individuals.

**Methods:** A total of 117 participants (69 women; 39.9 ± 11.4 years, mean ± std) underwent home-based polysomnography (PSG) and simultaneous WSA recording. Data analysis focused on evaluating sleep-wake distinction and sleep stage identification using standard classification metrics.

**Results:** WSA demonstrates high sensitivity (93%) for sleep detection and moderate sensitivity (73%) for wakefulness, achieving an overall accuracy of 87% for sleep-wake distinction. The device showed consistent performance across various demographic subgroups, including different age, BMI, mattress and sleep arrangements (with or without bed partner) categories. Challenges were noted in accurately classifying specific sleep stages, particularly in distinguishing between light and deep sleep, with a mean accuracy of 63% and a Cohen’s Kappa of 0.49. The WSA tended to overestimate total sleep time (+20 min) and light sleep (+1h21 min) while underestimating REM (−15 min) and deep sleep (−46 min) durations. Disagreements between expert reviewers, particularly between light and deep sleep stages, mirrored in part the WSA’s misclassifications. Participants reported significantly altered perceived sleep quality during the night with the PSG, suggesting potential discomfort during sleep.

**Conclusions:** WSA offers a promising approach to sleep monitoring in natural home environments. Being contactless and placed under the mattress, the WSA’s allows for long-term monitoring of sleep measures. It shows competitive performance in sleep-wake and sleep stage identification compared to other consumer devices. Progress in wearable and nearable devices is necessary to enhance their accuracy to better support the monitoring of populations with strongly impaired sleep, although limited by an imperfect gold standard. This work also emphasizes the importance of using large, diverse, and challenging datasets, as well as the need for a standardized methodology for accurate sleep stage classification.

## Introduction

Sleep is a vital pillar of human health, crucial for executing daily tasks and sustaining overall well-being [1]. Poor sleep quality can significantly impact life across various dimensions, including physical health, social interactions, and emotional stability [2,3]. Poor sleep quality diminishes alertness and cognitive function, increasing the risk of accidents. Furthermore, inadequate sleep is linked to numerous health issues, such as metabolic disorders, cardiovascular disease, neurocognitive impairments, and mental health challenges [4]. In severe cases, poor sleep quality can lead to critical outcomes, including an increased risk of suicide [5]. Despite its critical role, research shows that a significant portion of the population struggles with sleep-related issues. Approximately 30% of adults permanently experience at least one symptom of nocturnal insomnia [6], over 15% report sleepiness that affects daily activities [7], and more than 10% suffer from obstructive sleep apnea [8].

The ability to assess sleep architecture and the transitions between wakefulness and sleep from night to night is crucial for understanding sleep health. This information is captured in the hypnogram, a visual representation of the different sleep stages throughout the night, which can be obtained through polysomnography (PSG). The hypnogram represents the architecture of sleep, providing a visual depiction of the different sleep stages throughout the night: N1, N2, N3 and REM stages [9]. Polysomnography (PSG), which is considered the gold standard for identifying the micro and macro sleep architecture, records a variety of signals (electroencephalogram (EEG), electrooculogram (EOG), electromyogram (EMG), electrocardiogram (ECG), pulse oximetry, air flow and breathing effort [10]). As the hypnogram helps the physician identify abnormal sleep patterns, quantify sleep fragmentation and analyze sleep stage distribution, it is essential for diagnosing various sleep disorders, such as sleep-related breathing disorders, narcolepsy, hypersomnia, and disturbing sleep-related behaviors [11].

PSG, while a valuable tool, often involves extensive setup time and significant costs, making it less feasible for routine monitoring. The PSG is then scored manually, which is a labor-intensive process that can lead to long waiting times, and is prone to human error. Additionally, since it is typically conducted over just a few nights, typically with a gold standard of one night, it fails to capture the full picture of an individual’s circadian rhythms or night-to-night variability [12,13]. In most cases, PSG is conducted in a sleep lab with numerous sensors that can disrupt natural sleep patterns For all these reasons, PSG results in data that may not accurately represent a person’s typical sleep in their own home [14], and therefore lacks ecological validity. Home-based sleep monitoring devices offer a more ecologically valid approach, as they allow for continuous sleep tracking in the comfort of one’s own home. This provides a more realistic picture of an individual’s sleep patterns and helps to address the limitations of traditional sleep studies. The need for more accessible and cost-effective solutions for continuous, multi-night sleep monitoring is driven by the desire to enhance our understanding of sleep quality and facilitate the identification of potential sleep disorders.

To address the challenges of cost and complexity in multi-night assessments, wearable sleep trackers are commonly used to detect sleep and wake states through body movements, using actigraphy-based devices [14]. These devices, while often marketed as a solution for sleep monitoring, still show two limitations. Firstly, they struggle to accurately differentiate between wakefulness and sleep when individuals remain still while awake [15]. Secondly, wearable devices also require users to maintain proper charging and wearing, leading to frequent data loss and inconsistent performance [16]. In contrast, nearables, which are not in direct contact with the individual, eliminate the need for charging or wearing a device. Generally positioned under the mattress or at the bedside, they require minimal intervention after setup.

Withings Sleep Analyzer (WSA) is a nearable device offering a promising non-invasive alternative for monitoring sleep stages over consecutive nights. Despite the proliferation of wearable and nearable sleep-tracking devices, there is a notable lack of rigorous evaluation of the algorithms, leading to concerns about data accuracy and reliability [17,18]. In contrast, the WSA performance to detect apnea was validated in the laboratory [19–23], for which it is CE marked and FDA-cleared. In addition, it was assessed for nocturnal sleep periods detection [24] and cardiorespiratory monitoring [25]. A study evaluating sleep stage detection by WSA is still lacking. Our work aims to assess the WSA’s sleep stage identification algorithm in a real-world home setting with 117 healthy participants.

There are several important methodological issues we aim to address in this work. Existing research in sleep-tracking technologies highlights several persistent problems and limitations. Many studies, such as those on the WatchPAT [26] and Somnofy [27] devices, demonstrate potential but often lack comprehensive validation across diverse populations and real-world settings. Evaluations of consumer-grade devices, like those conducted by Chinoy et al. [28] and Kainec et al. [29], are frequently limited by small sample sizes and strict exclusion criteria (young individuals with controlled caffeine, alcohol, screen time, and sleep regularity), which limit the generalizability of their findings. These studies typically occur in controlled environments, such as sleep labs, which do not accurately reflect regular user conditions at home.

Moreover, the absence of synchronized epoch-by-epoch data and reliance on lights-off and lights-on limits in these studies further complicate the accuracy assessments of sleep stage detection. This lack of detailed, labeled data can lead to discrepancies in sleep onset and offset times, affecting the reliability of the results.

Recent publications emphasize the importance of assessing these devices under everyday conditions, aligning with our work, eg, [30]. Other works emphasize the need for standardized evaluation protocols and methodological consistency across studies, reinforcing the approach taken in our research [31,32]. These studies collectively highlight the critical need for standardized and rigorous evaluation methods in sleep monitoring technology. Benchmarking efforts of devices are also being addressed, as recent studies manifest [28,29,33].

Our study uniquely contributes by assessing the WSA’s performance in a home setting, on a large and diverse sample, in free living conditions, that is without constraints about substance intake, activity before bed, or forced sleeping period, providing insights into the device’s accuracy and reliability in real-world conditions. The primary goal is to evaluate WSA’s performance using classification metrics and mean absolute error (MAE) on standard sleep metrics [9]. Additionally, we aim to compare performance across subgroups based on demographic and clinical characteristics, such as sex, age, BMI, bed partner presence, mattress type, but also sleep quality Pittsburgh Sleep Quality Index (PSQI) score [34], mattress and thickness.

By addressing these gaps, our study seeks to enhance the understanding of WSA’s capabilities and contribute to the development of more reliable and user-friendly sleep-tracking technologies.

## Materials and Methods

### Dataset

#### Participants

Data were collected during a clinical study that was approved by the French Institutional Review Board (Approval Reference Number: 2018-A03129-46) and conducted by Withings from 2020 to 2022. The inclusion process for the study was conducted by a specialized recruitment company. The cohort consisted of 194 volontary individuals from the general population, who did not have any pre-existing medical conditions that would necessitate a PSG examination. The recruitment process aimed to ensure a balanced distribution of BMI within each age group.

A total of 194 participants slept for the first night at home with Withings Sleep Analyzer (WSA), and the CID-LXe portable PSG (CIDELEC, Ste Gemmes sur Loire, France), both installed by a technician the evening before the recording. The night after that, the participants slept at home with the Withings Sleep Analyzer, without the PSG. Each participant was asked to answer once to the Pittsburgh Sleep Quality Index (PSQI) questionnaire upon enrolment. After each night, participants were asked to complete a survey addressing the following: their sleep quality for that night, a comparison of that night’s sleep quality to their usual sleep, whether they took sleeping pills, and any sleep disturbances caused by the sensors.

As illustrated in the flowchart (Figure 1), several sources of data loss have led to the inclusion of 117 participants in this analysis. This trial has been particularly affected by data collection issues due to a dedicated firmware meant to collect high frequency sensor data (reason for exclusion A).

**Figure 1:**
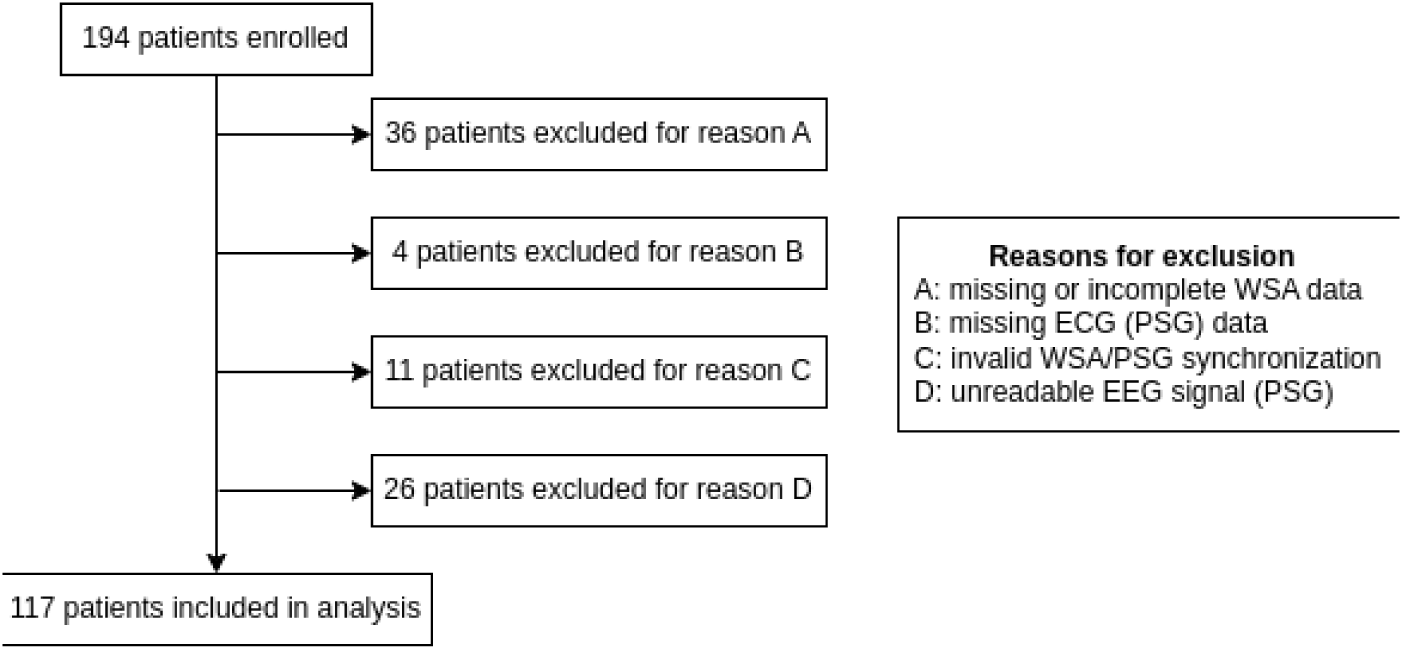
Study inclusions flowchart.

#### Procedure/Study Design

WSA sampled the raw data from the pressure and microphone sensors (respectively 250 Hz and 8 Hz). The reference device was CID-LXe PSG that recorded the following signals: 3 EEG, 1 ECG, 2 EOG, 1 submental EMG and ambient light. Based on experience from past investigations, the digital oximeter and the nasal cannula are the major sleep disruptive sensors during PSG, even in a home setting. To minimize this sleep disruption and obtain the most natural possible sleep, PSG was therefore done at home with only EEG, EMG, ECG and ambient light sensors. As sleep stages annotations from expert reviewers are based on EEG and lights sensors only [35], the gold standard reference quality is not affected by the removal of the oximeter and the nasal cannula.

### Data Processing

WSA is designed to identify sleep stages over 6-minute windows. As a result, this analysis focuses on evaluating WSA’s performance in detecting stages longer than 6 minutes. It should be kept in mind that sleep stages shorter than this are ignored by the device and in this study. In addition, WSA does not distinguish stages N1 and N2, so for the aim of comparison, the consensus sleep stages N1 and N2 obtained from the PSG were combined into the “light” category. REM and awake stages of the WSA and the consensus are directly comparable, and N3 is compared with the stage “deep sleep” from the WSA. The resulting stages are illustrated in Figure 2.

**Figure 2:**
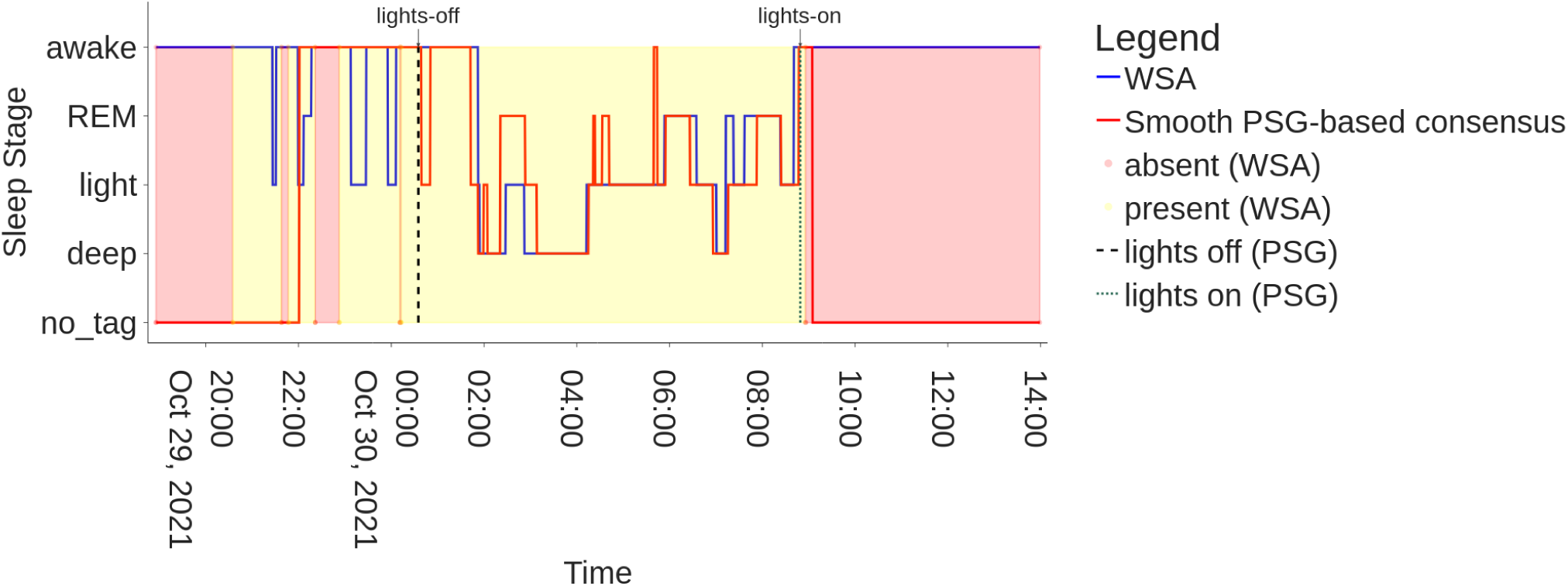
Example of hypnogram (subject 20211029_1) showing WSA sleep stages and the reference (PSG consensus)

Two independent certified sleep technologists annotated the PSG signals in 30-second epochs, classifying sleep stages as N1, N2, N3, REM, and awake, following the AASM scoring manual [9]. A third professional reviewed and established a consensus for reference data. Sleep stage comparisons are performed over windows of 30 seconds. To evaluate WSA’s performance in detecting sleep stages longer than 6 minutes, consensus data are smoothed by selecting for each 6-minute window the stage with the highest occurrence of 30-second epochs. For more details about the procedure, please refer to Supplementary Materials A.

In addition to standard sleep quality measures calculated over the night, such as total sleep time and the duration of each stage, evaluating stage classification requires synchronizing epoch-by-epoch data from both devices over time. Synchronization of the clocks of the devices was obtained by maximizing the correlation of the breathing signals of the two devices [36]. Details of the procedure can be found in Supplementary Materials A.

An example hypnogram is presented on Figure 2, showing the smoothed, PSG-based consensus in red and WSA sleep stages in blue. WSA indicates bed absence/presence, marked respectively by a red and yellow background, information which is absent from the PSG data. The consensus annotations use instead the events lights-on/off detected by the ambient light sensor of the PSG as proxies for bed in and bed out events. For further information on challenges arising from using lights-off/on timestamps as proxies for bed presence, please refer to Supplementary Materials A.

### Performance Evaluation Metrics and Tools

#### Sleep Quality Measures

Standard sleep quality measures utilized for this analysis and based on the AASM scoring manual [9] are the following : total sleep time (TST), time spent in each sleep stages (TIREM, TILight and TIDeep), proportion of the time spent in each sleep stage compared to total sleep time (PIREM, PILight, PIDeep) and number of episodes in each sleep stage (NEREM, NELight and NEDeep). Each of these measures have been computed by each device (PSG and WSA) and then compared using the Mean Absolute Error (MAE) and a Bland Altman analysis, described in Supplementary Materials B.

#### Epoch-by-epoch Classification Evaluation

We assess the classification of sleep and wake phases, and the classification of sleep stages. Standard classification metrics are used in this work: accuracy, kappa, sensitivity, specificity.

#### Subgroup Analyses

Subgroup analyses were conducted to determine whether WSA performance is affected by the following factors: 1) demographic: age, BMI, sex, 2) sleep quality factors: PSQI score, PSQI components, hypnotics, reported sleep quality, and whether the participant has woken up at least once during the night of the recording, 3) environmental: whether the subject has a bed partner, mattress type, mattress thickness.

#### Boxplots by metadata categories

Boxplots summarize the distribution of a dataset by displaying its median, quartiles, and potential outliers. They are useful in this context because they provide a clear visual comparison of the performance across different population subgroups.

#### Statistical Tests

To compare the median MAE between different groups (categories of a given metadata as listed in Table C.1), a Kruskal-Wallis test has been performed on the MAE obtained on all the metadata described in Table C.1 for each of the following sleep quality measures (TST, TIREM, TILight, TIDeep, PIREM, PILight and PIDeep) but also for the number of episodes in each sleep stage. To compare the MAE variance between different groups, a Brown-Forsythe test has been performed on the same data for the same sleep quality measures. The Holm-Bonferroni correction is employed after both statistical tests to address the issue of multiple comparisons.

## Results

This section assesses the Withings Sleep Analyzer (WSA) against polysomnography (PSG) for sleep monitoring. Among 117 healthy participants, the WSA showed high sensitivity (93 %) and moderate specificity (73 %) for sleep detection, with an overall accuracy of 87% for sleep-wake distinction. Challenges persist in distinguishing light from deep sleep, similar to issues faced by expert reviewers. The WSA performed consistently across demographic subgroups. Additionally, wearing the PSG sensors was associated with a degraded sleep quality. These findings affirm the WSA’s reliability while highlighting areas for improvement in sleep stage differentiation. The WSA did not impact user comfort.

### Patient characteristics

Demographic and clinical data are summarized in Table 1 for the population included in the analyses.

**Table 1:** Demographic characteristic and clinical data for the entire population. For quantitative data, results are presented as mean (SD). SD: standard deviation.

| Demographics | Age (years) | Height (cm) | Weight (kg) | BMI (kg/m<sup>2</sup>) | Male | Female | Skin type 1* | Skin type 2* | Skin type 3* |
| --- | --- | --- | --- | --- | --- | --- | --- | --- | --- |
| Mean (SD)<br>or<br># and proportion (%) | 39.9 (11.4) | 171.6 (8.3) | 70.4 (12.0) | 23.9 (3.3) | 48 (41 %) | 69 (59 %) | 85 (75 %) | 21 (18 %) | 8 (7 %) |
| Min / Max | 19 / 69 | 153 / 194 | 49 / 103 | 17.9 / 30.9 | - |  |  |  |  |
| Known pathologies | Asthma | Arterial hypertension | Bipolar disorder | Ulcerative colitis | Ankylosing spondyl |  |  |  |  |
| Nb (%) | 5 (4 %) | 3 (< 3 %) | 1 (< 1 %) | 1 (< 1 %) | 1 (< 1 %) |  |  |  |  |
| Pathologies | Cardiovascular<sup>1</sup> | Neurological<sup>2</sup> | Digestive<sup>3</sup> | Metabolic<sup>4</sup> | Psychiatric<sup>5</sup> | Other |  |  |  |
| Nb (%) | 4 (4 %) | 0 (0 %) | 0 (0 %) | 2 (2 %) | 0 (0 %) | 6 (5%) |  |  |  |
| Pittsburgh Sleep Quality Index questionnaire [34] | PSQI score | Component 1:<br>Subjective sleep quality | Component 2:<br>Sleep latency | Component 3:<br>Sleep duration | Component 4:<br>Habitual sleep efficiency | Component 5:<br>Sleep disturbances | Component 6:<br>Use of sleeping medications | Component 7:<br>Daytime dysfunction |  |
| Mean (SD) | 6.0 (2.9) | 1.2 (0.7) | 1.3 (0.9) | 0.8 (0.8) | 0.4 (0.7) | 1.3 (0.5) | 0.2 (0.6) | 0.9 (0.8) |  |
| Min / Max | 0 / 15 | 0 / 3 | 0 / 3 | 0 / 2 | 0 / 3 | 0 / 3 | 0 / 3 | 0 / 3 |  |
| Medications | Antihypertensive | Antidepressant | Hypnotics | Corticoid | Antiepileptic | Antipsychotic |  |  |  |
| Nb (%) | 3 (3 %) | 2 (2 %) | 1 (1 %) | 1 (1 %) | 1 (1 %) | 1 (1 %) |  |  |  |
| Mattress type | foam | spring | latex | memory foam | other | Unknown |  |  |  |
| Nb (%) | 41 (35 %) | 29 (25 %) | 24 (21 %) | 12 (10 %) | 8 (7 %) | 3 (3 %) |  |  |  |
| Bed partner | Yes | No |  |  |  |  |  |  |  |
| Nb (%) | 64 (55 %) | 52 (45 %) |  |  | *Skin type is based on reduced Fitzpatrick scale<br><sup>1</sup> (arterial hypertension, atrial fibrillation, myocardial infarction)<br><sup>2</sup> (Parkinson's, stroke, adenoma, manic-depression, epilepsy)<br><sup>3</sup> (hemolytic anemia due to glutathione reductase deficiency, ulcer, sleeve, diverticula)<br><sup>4</sup> (diabetes, hypothyroidism, dyslipidemia, obesity/overweight)<br><sup>5</sup> (depression, insomnia, anxiety) |  |  |  |  |
| Sensors disturbance (due to PSG device) | Yes | No |  |  |  |  |  |  |  |
| Nb (%) | 46 (39 %) | 71 (61 %) |  |  |  |  |  |  |  |

A total of 117 participants were included in the analysis. The cohort comprised 60 % women and 40 % men, with a mean age of 39.9 years (SD = 11.4), ranging from 19 to 69 years. The average BMI was 23.9 kg/m² (SD = 3.3).

Regarding known pathologies, most of the participants were healthy. Still, 4 % of participants reported asthma, while less than 3% reported arterial hypertension. Other conditions such as bipolar disorder, ulcerative colitis, and ankylosing spondylitis were present in less than 1 % of the cohort. Pathologies were relatively rare, as participants were supposed to be healthy.

The average sleep quality in this population is considered mildly impaired, with a mean Pittsburgh Sleep Quality Index (PSQI) score of 6 out of 21. The standard deviation of 2.9 highlights significant variability. This means that a portion of the population might experience quite good sleep quality (scores 0-5), while others might have moderate sleep difficulties (scores 8-14).

The most common mattress types were foam (35%), spring (25%), and latex (21%). Fifty-five percent of participants reported having a bed partner. Sensor disturbances were reported by 46 participants (39 %) due to the PSG device, while no disturbances were reported regarding the WSA nearable.

### Sleep Stages Classification

Figure 3 presents the confusion matrix containing the epochs across all nights, and Table 2 gathers sleep stages classification results on the whole dataset.

**Figure 3:**
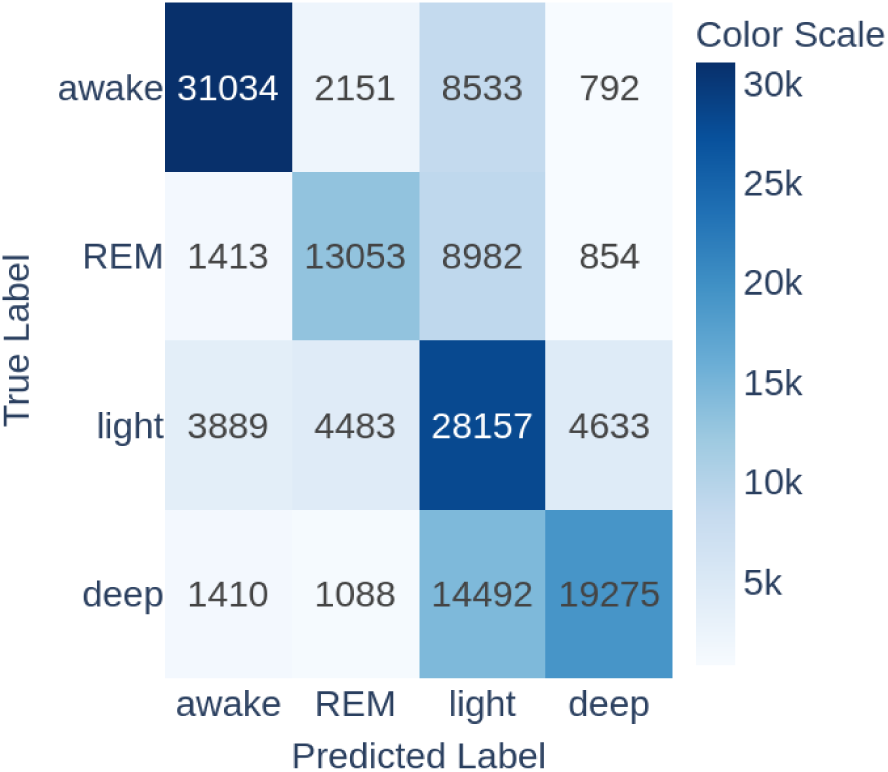
Sleep stage classification confusion matrix (WSA sleep stage detection vs. PSG ground truth)

**Table 2:** Summary of sleep stage classification results over nights (computed as the averages of all nights)

| Mean of results per night | Sleep stages |  |  |  |  |  |  |  | Sleep-Wake |  |  |  |
| --- | --- | --- | --- | --- | --- | --- | --- | --- | --- | --- | --- | --- |
|  | Kappa | Accuracy | Se REM | Se light | Se deep | Sp REM | Sp light | Sp deep | Kappa | Accuracy | Se sleep | Sp sleep |
| Mean | 0.49 | 0.63 | 0.53 | 0.68 | 0.56 | 0.93 | 0.69 | 0.94 | 0.65 | 0.87 | 0.93 | 0.73 |
| 95% CI | [0.47, 0.51] | [0.62, 0.65] | [0.48, 0.57] | [0.65, 0.71] | [0.51, 0.60] | [0.92, 0.94] | [0.67, 0.71] | [0.93, 0.95] | [0.62, 0.69] | [0.86, 0.89] | [0.92, 0.94] | [0.69, 0.77] |
| SD | 0.12 | 0.09 | 0.24 | 0.16 | 0.25 | 0.05 | 0.11 | 0.06 | 0.19 | 0.07 | 0.06 | 0.22 |

#### Sensitivity and Specificity

The WSA demonstrated a mean sensitivity of 93 % for detecting sleep, meaning it correctly identifies when a person is asleep 93 % of the time. The WSA showed a mean specificity of 73 %, indicating it accurately detects when a person is awake 73% of the time.

The performance in accurately classifying each individual sleep stage against the other three is more variable, with sensitivities of 53 %, 68 %, and 56 % for REM, light sleep, and deep sleep respectively. The specificities were 93 %, 69 %, and 94 % for REM, light and deep sleep respectively.

#### Cohen’s Kappa and Accuracy

Please note that contrary to sensitivity and specificity, Cohen’s Kappa and accuracy depend on the relative sizes (prevalences) of each class. In this study’s dataset, the WSA achieved a mean Cohen’s Kappa of 0.65 for distinguishing between sleep and wake stages, and 0.49 for differentiating among the four stages. WSA accuracy to distinguish between sleep and wake was 87 %. The mean accuracy for the 4 stages was 63 %.

### Assessment of Sleep Quantity Measures

This section explores the agreement between WSA and PSG for quantitative sleep measures, using regression analysis, the mean absolute error, and Bland-Altman analysis.

As shown in Figure 4, WSA demonstrated a slope estimate for total sleep time (TST) of 0.98 (by the RANSAC algorithm, see Supplement materials B), which was statistically significant (p < 0.05). The R-squared value for TST was 0.89 with 38 % of outliers, indicating a moderate linear relationship with PSG.

**Figure 4:**
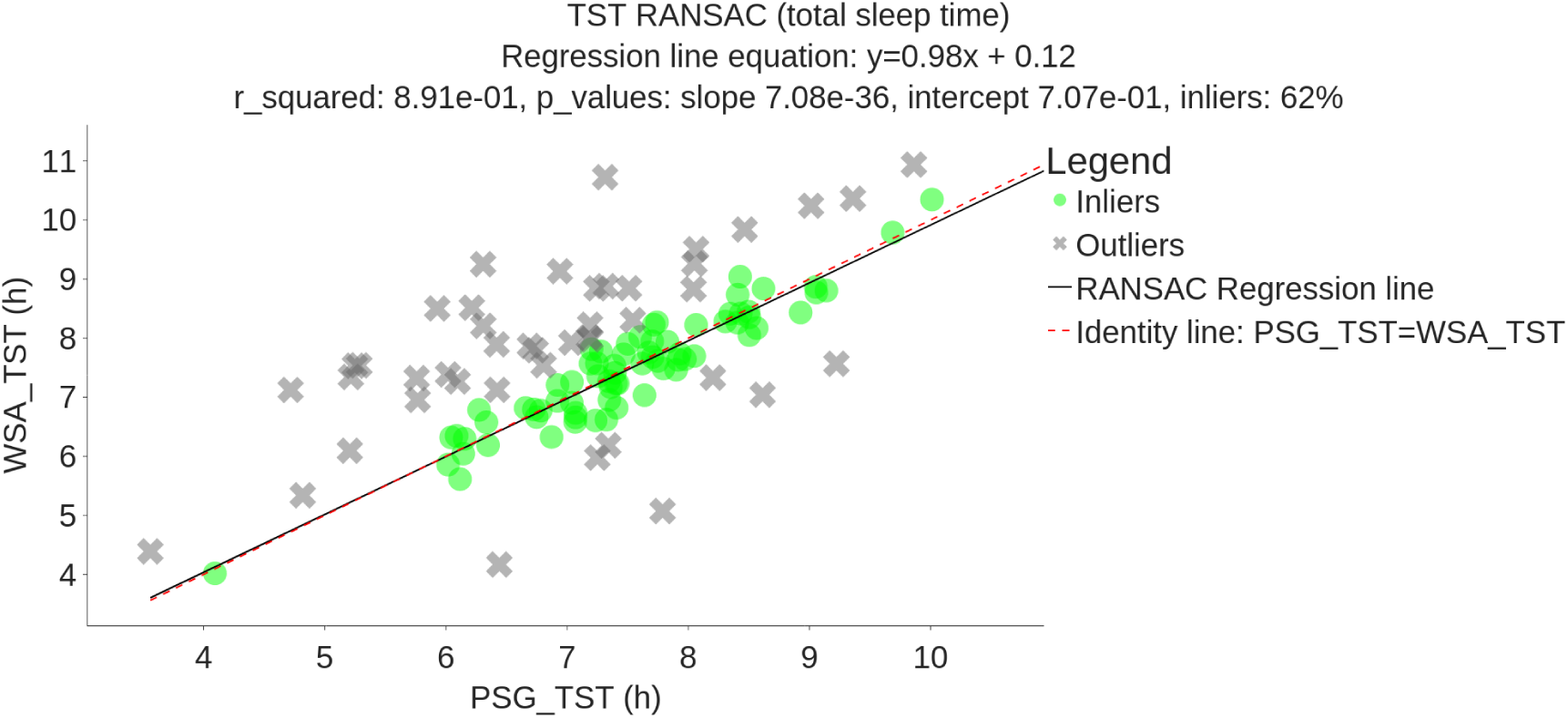
RANSAC Regression plot between WSA and PSG total sleep time (black line) compared to identity line (red dashed line) on 117 subjects (r² = 0.89, slope p-value: 7e-36, inliers: 62 %)

WSA was not correlated to time spent in deep sleep (slope estimate of 0.05, p = 0.7, Figure D3). For light sleep time, the regression estimate was 0.4, with statistical significance (p < 0.05, Figure D2). REM sleep time had a regression estimate of 0.7, also statistically significant (p < 0.05, Figure D1).

Mean absolute error (MAE) is used to evaluate the precision with which the WSA estimates standard sleep quality measures, taking the PSG-based consensus as “the truth”. The MAE values of total sleep time, REM duration, light sleep duration and deep sleep duration are respectively 46 min, 35 min, 1 h 33 min and 1 h 05 min, with average PSG values of respectively 7 h 15 min, 1 h 43 min, 2 h 57 min, and 2 h 35 min. The average number of episodes in each sleep stage and the average number of stage changes can be found in Table 3. The WSA tends to underestimate the number of episodes for each sleep stage and is less sensitive to transitions between epochs.

**Table 3:**
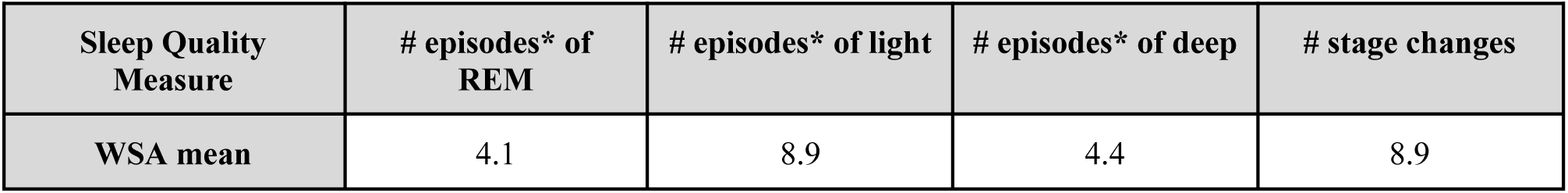
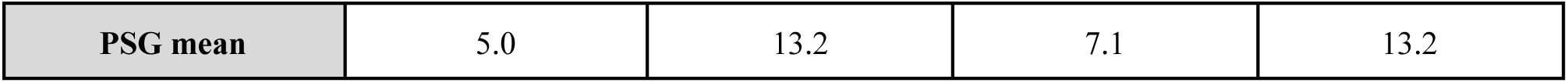
Comparison of Sleep Stage Episodes and Stage Changes Between WSA and PSG on 117 subjects. *An episode as a sequence of consecutive epochs in the same sleep stage.

The results of the regressions are summarized in Table 4. Additional scatter plots and regression analyses of these metrics can be found in Supplementary Materials D.

**Table 4:**
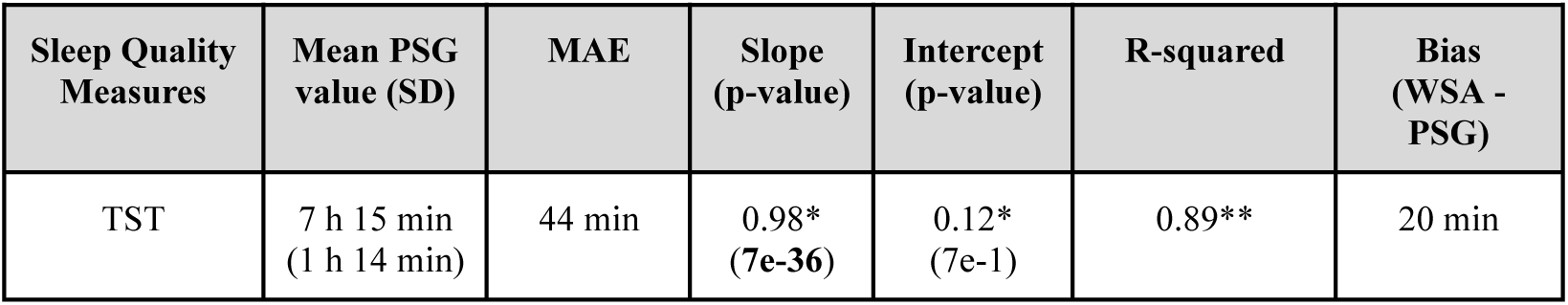
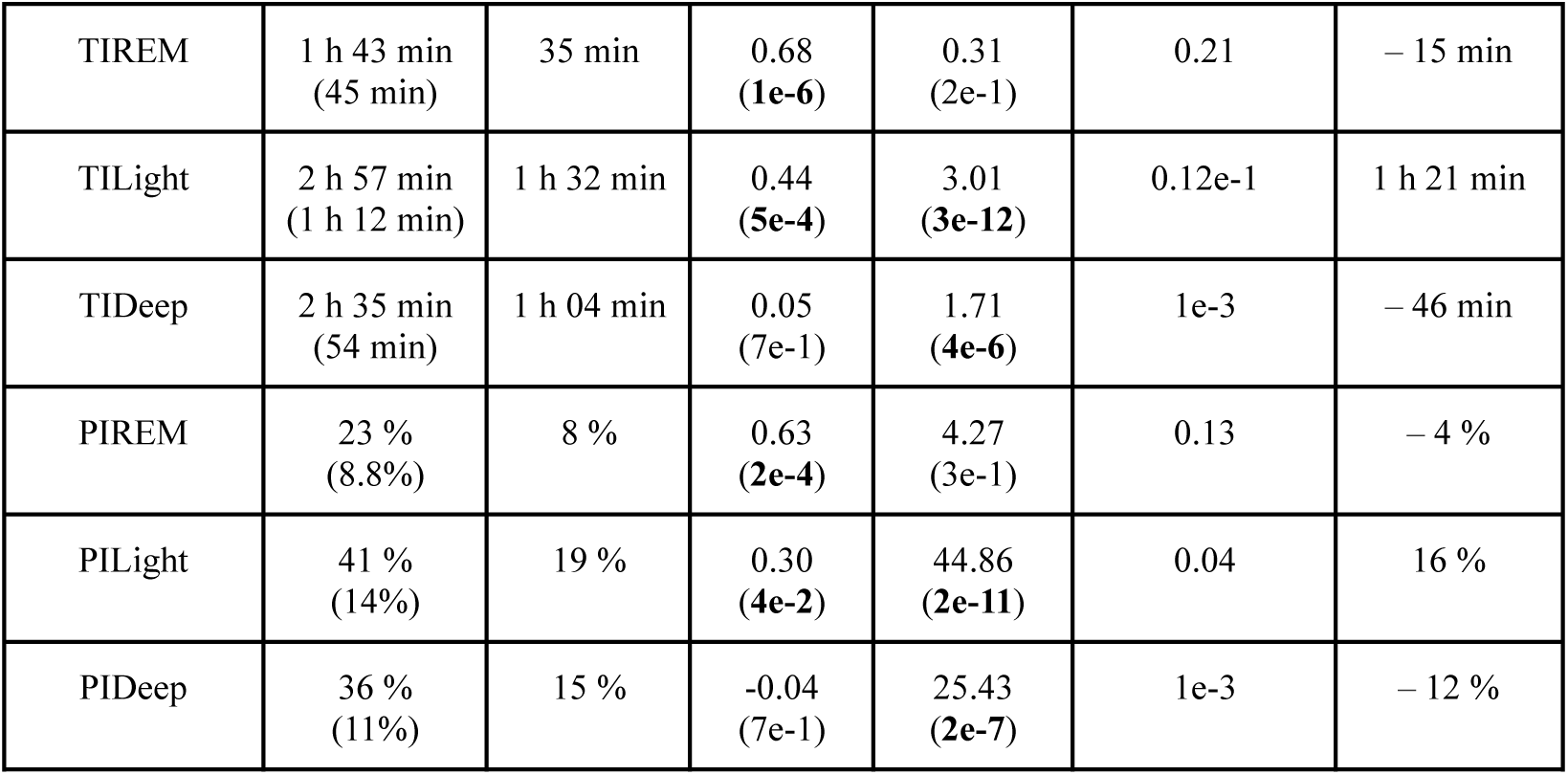
Summary of regression and Bland-Altman results obtained on sleep-related metrics between WSA and PSG estimations. *Results obtained with RANSAC method. Other regression results are obtained using the Ordinary least squares (OLS) method. **Bold p-values**: p < 0.05

| Sleep Quality Measures | Mean PSG value (SD) | MAE | Slope (p-value) | Intercept (p-value) | R-squared | Bias (WSA - PSG) |
| --- | --- | --- | --- | --- | --- | --- |
| TST | 7 h 15 min<br>(1 h 14 min) | 44 min | 0.98*<br>(7e-36) | 0.12*<br>(7e-1) | 0.89** | 20 min |
| TIREM | 1 h 43 min<br>(45 min) | 35 min | 0.68<br>**(1e-6)** | 0.31<br>(2e-1) | 0.21 | – 15 min |
| TILight | 2 h 57 min<br>(1 h 12 min) | 1 h 32 min | 0.44<br>**(5e-4)** | 3.01<br>**(3e-12)** | 0.12e-1 | 1 h 21 min |
| TIDeep | 2 h 35 min<br>(54 min) | 1 h 04 min | 0.05<br>(7e-1) | 1.71<br>**(4e-6)** | 1e-3 | – 46 min |
| PIREM | 23 %<br>(8.8%) | 8 % | 0.63<br>**(2e-4)** | 4.27<br>(3e-1) | 0.13 | – 4 % |
| PILight | 41 %<br>(14%) | 19 % | 0.30<br>**(4e-2)** | 44.86<br>**(2e-11)** | 0.04 | 16 % |
| PIDeep | 36 %<br>(11%) | 15 % | -0.04<br>(7e-1) | 25.43<br>**(2e-7)** | 1e-3 | – 12 % |

The Bland-Altman plots are presented in Figures 5 and 6. Biases are summarized in Table 4.

**Figure 5:**
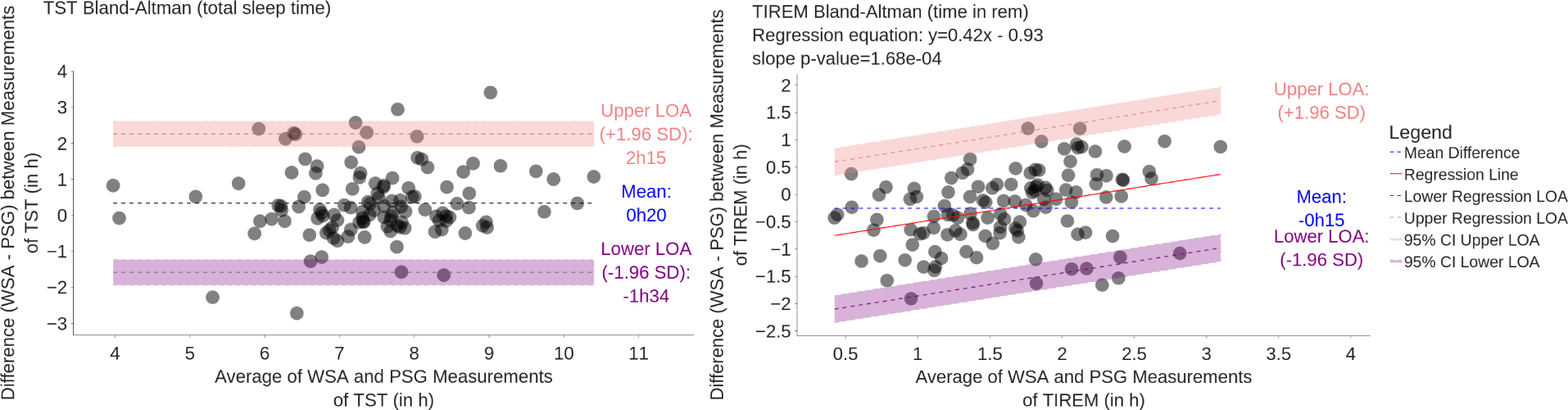
Bland-Altman Plot Comparing WSA and PSG Estimation of total sleep time and time in REM on 117 subjects.

**Figure 6:**
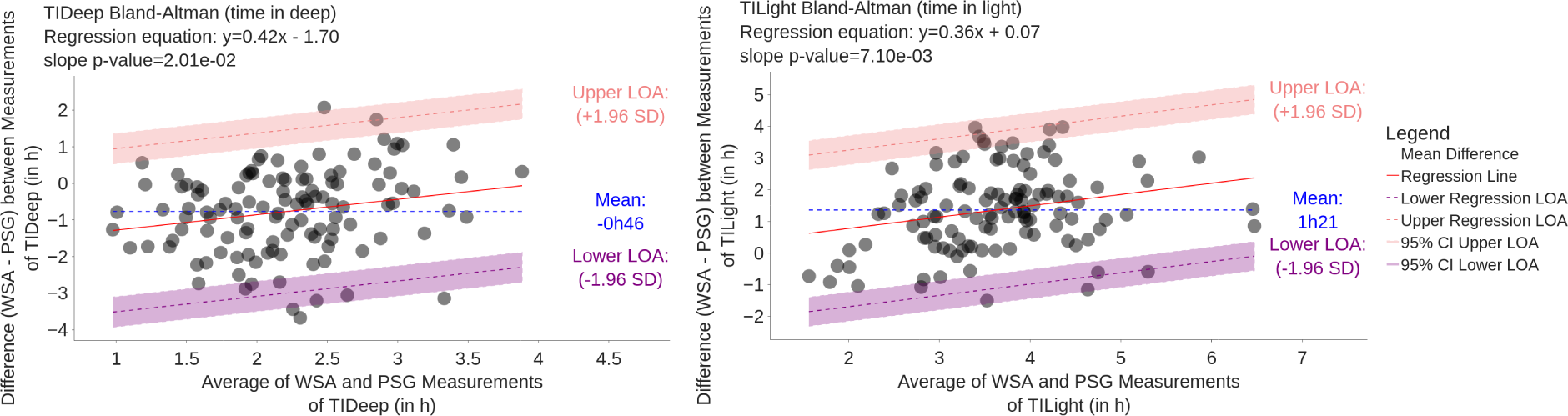
Bland-Altman Plot Comparing WSA and PSG Estimation of time in deep and time in light on 117 subjects.

On Figure 5, the Bland-Altman plot for TST does not reveal any tendency with the duration of the night. The mean error is 20 minutes. The wide limits of agreement show that there is a substantial amount of variability between PSG and WSA. By contrast, the average error in REM, light and deep sleep durations depend on the duration of the phase. Light sleep is *on average* overestimated for any duration, deep sleep is *on average* underestimated for any duration, and REM is *on average* underestimated for short and overestimated for long durations.

The mean error (bias) in light sleep duration is important, with 1h 21 min. Biases in REM and deep sleep are respectively −15 min and −46 min. The mean error in TST is therefore 20 min.

### Subgroup Analyses

No significant differences were found in the median or variance of the error across most groups studied for all the quantitative sleep variables of Table 4. This includes comparisons based on gender, BMI categories, age groups, sleeping arrangements, PSQI scores, self-reported sleep quality, mattress thickness, and night awakenings. However, an exception was noted in the median proportion of deep sleep, where significant differences were observed within the mattress type subgroup with a median MAE of respectively 12%, 22%, 11% and 7.4% for spring, foam, memory foam, and latex. This finding suggests that mattress type may influence the measurement of deep sleep, highlighting an area for further investigation. For a comprehensive overview of the statistical tests and detailed results, please refer to the Supplementary Materials E.

As an illustration, Figures 7 to 9 show the boxplots of TST mean absolute error of WSA compared to PSG, stratified by age groups, BMI ranges, and whether the subject was alone in bed. They suggest that the WSA performance did not depend on these factors. Boxplots for TST MAE for the other subgroups can be found in the Supplementary Materials D.

**Figure 7:**
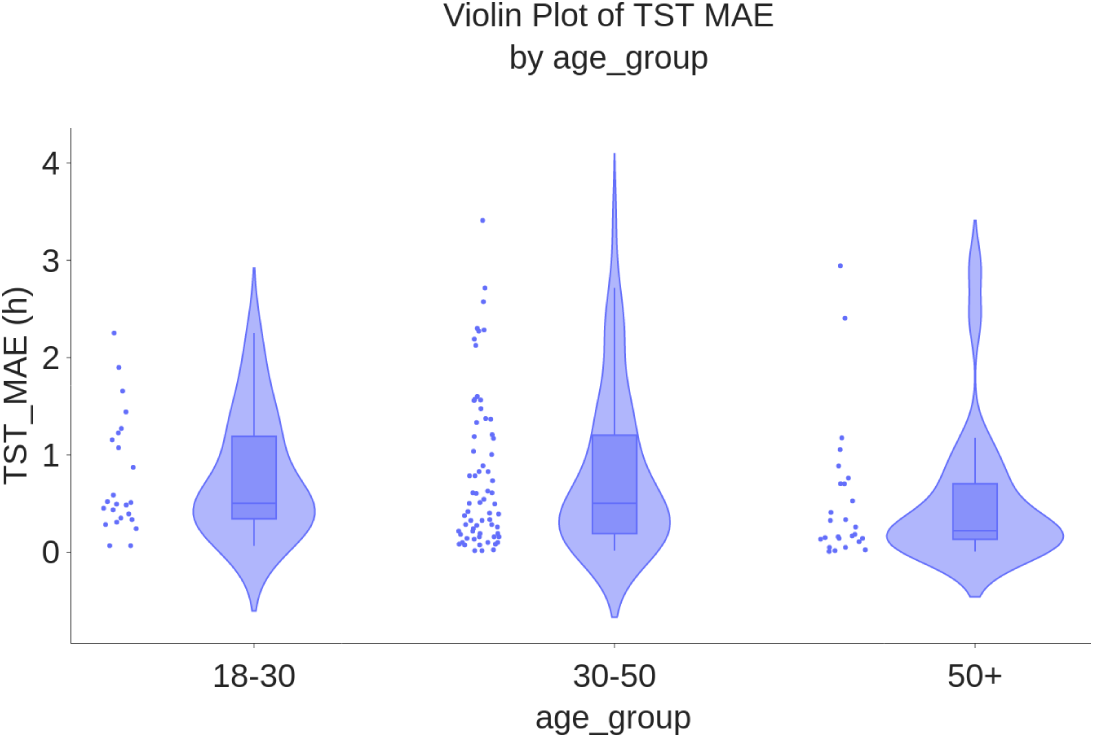
Boxplot of TST MAE (h) by age group on 117 subjects.

**Figure 8:**
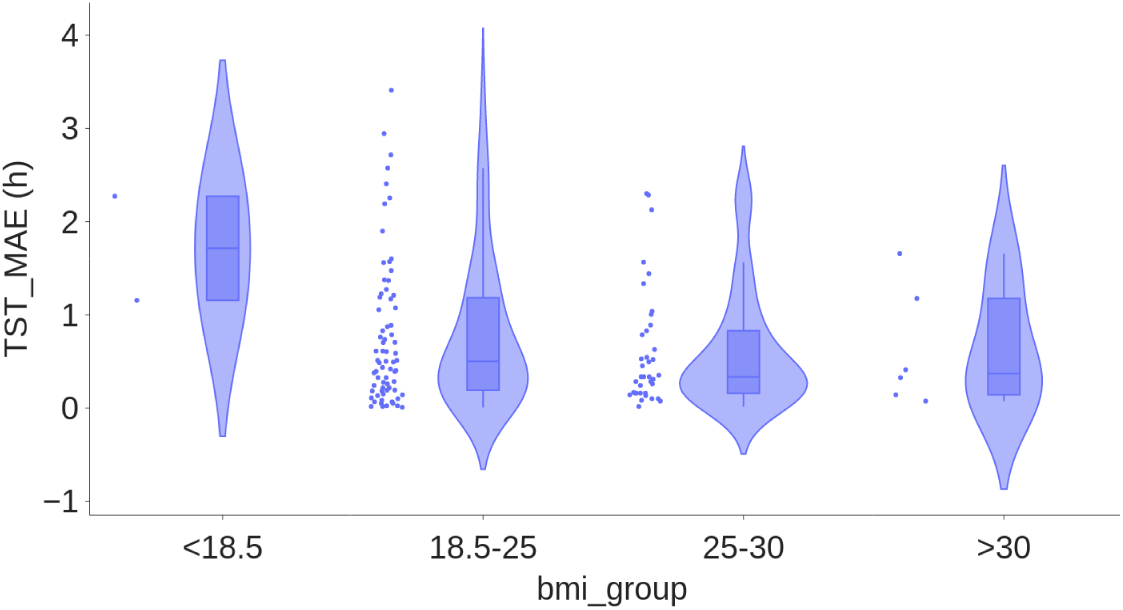
Boxplot of total sleep time MAE (h) by bmi group on 117 subjects.

**Figure 9:**
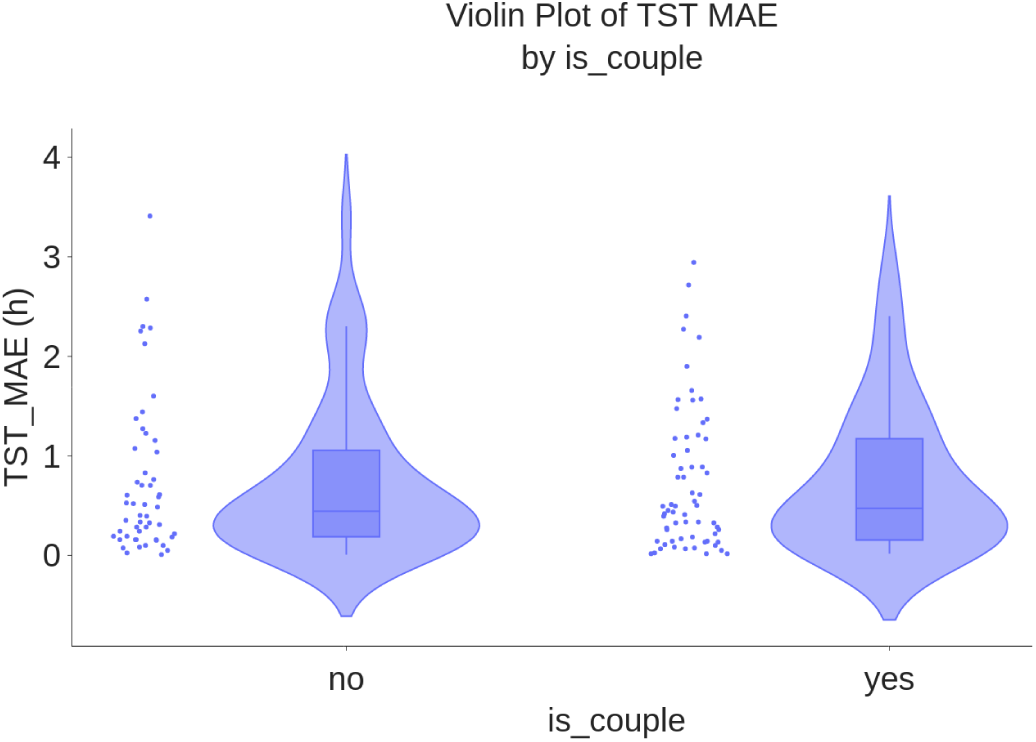
Boxplot of total sleep time MAE (h) by is_couple groups on 117 subjects no: subjects who slept alone the night of the recording yes: subjects who had a sleep partner the night of the recording.

**Figure 10:**
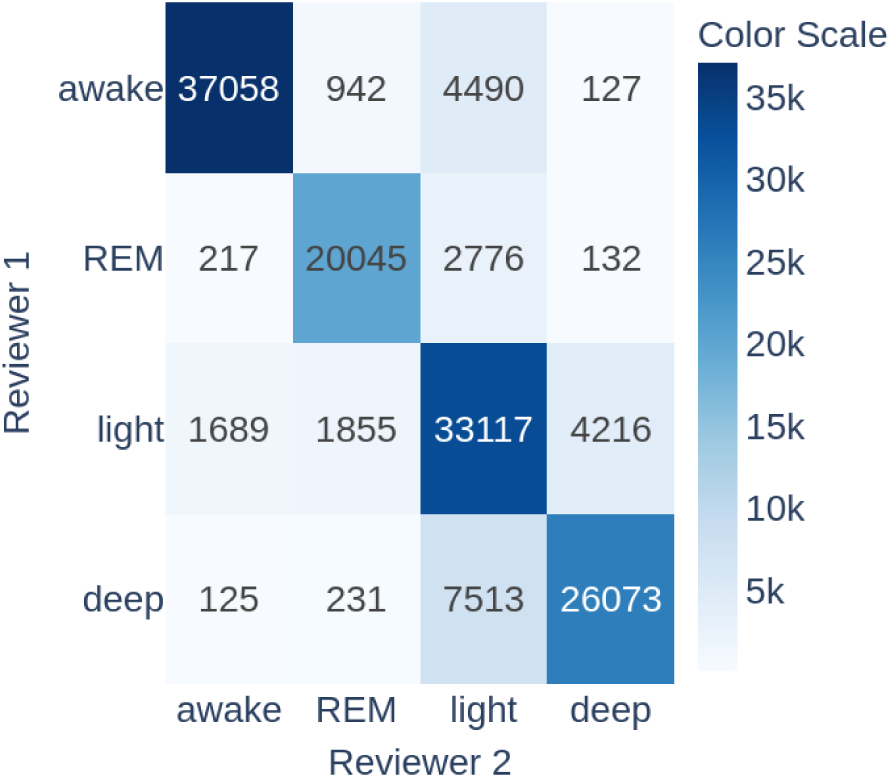
Sleep stage classification confusion matrix between first two reviewers annotations.

### Agreement Between Reviewers

Each 30-second epoch of the PSG data was independently annotated by two reviewers. The mean kappa value for sleep stage determination between reviewers before reaching consensus is 0.71 (SD = 0.11), closely aligning with literature findings (kappa = 0.76 in [37]), while it is 0.81 (SD = 0.17) for sleep-wake classification.

In the analysis of reviewer agreement on sleep stage classifications, it was observed that the primary disagreements occur between light and deep stages, as well as between light sleep and awake/REM stages. This indicates that even expert reviewers, considered the gold standard, face challenges in accurately distinguishing these stages due to their inherent similarities and subtle physiological differences. This inherent difficulty in distinguishing light, deep, awake, and REM sleep stages by human experts is reflected in WSA which was trained on PSG data labeled using a similar protocol. It is likely that the imprecision of the gold standard passed on to the device.

#### Discrepancies in Performance: Epochs with Reviewers Agreement versus Disagreement

Given the discrepancies encountered in PSG annotation among reviewers, it is pertinent to investigate whether the WSA performed better on epochs where reviewers reached consensus compared to those where disagreements were present. To perform this analysis, the dataset was divided into two groups of epochs: those where reviewers reached agreement and those where disagreements occurred. For the sleep-stage agreement, an epoch was marked as “agreement” if both reviewers assigned the same sleep stage, and “disagreement” otherwise. Similarly, for the Sleep-Wake (S-W) agreement, an epoch was considered “agreement” if both reviewers classified it as either sleep or wake, and “disagreement” otherwise. Separate performance analyses were then conducted on these two complementary groups.

Results are summarized in Table 5. In the *Agreement* column (resp. *Disagreement* column), only epochs where both reviewers agreed (resp. disagreed) were included, while epochs with disagreement (resp. agreement) were excluded. Figure 11 shows the confusion matrices obtained for both sets.

**Figure 11:**
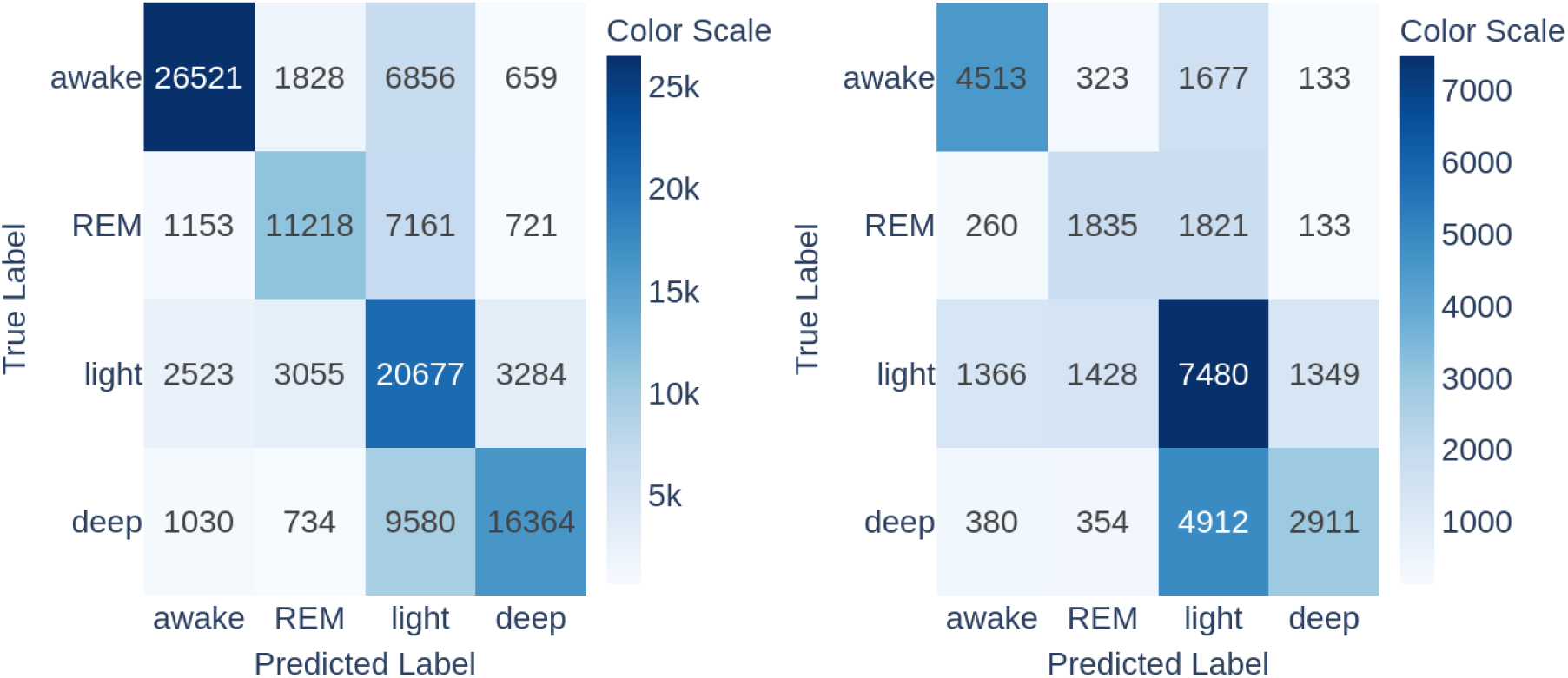
Sleep stage classification confusion matrices on sets with agreement (left) and disagreement (right) (WSA sleep stage detection vs. PSG ground truth)

**Table 5:** Summary of performance in reviewers agreement and disagreement subgroups.

| WSA vs. PSG |  | Agreement | Disagreement | Z score |
| --- | --- | --- | --- | --- |
| Nb epochs<br>(proportion of total) |  | 113,364<br>(79 %) | 30,875<br>(21 %) | - |
| Sleep Stages | Kappa | 0.52 | 0.27 | 78.06 |
|  | Accuracy (%) | 0.66 | 0.54 | 38.81 |
|  | Se REM | 0.55 | 0.45 | 31.21 |
|  | Se light | 0.7 | 0.64 | 20.16 |
|  | Se deep | 0.59 | 0.34 | 78.10 |
|  | Sp REM | 0.94 | 0.92 | 12.70 |
|  | Sp light | 0.72 | 0.56 | 53.69 |
|  | Sp deep | 0.95 | 0.93 | 13.75 |
| Nb epochs<br>(proportion of total) |  | 133,447<br>(93 %) | 10,792<br>(7 %) | - |
| Sleep-Wake | Kappa | 0.62 | 0.33 | 59.11 |
|  | Accuracy (%) | 0.88 | 0.76 | 35.77 |
|  | Se wake | 0.74 | 0.69 | 11.34 |
|  | Se sleep | 0.94 | 0.86 | 32.20 |

We used a Z-test for proportions to test for differences between sensitivity, specificity, accuracy, and kappa. All the results are statistically significant, WSA performing better on epochs where the two reviewers agreed. The progression is particularly significant for the sensitivity in deep sleep, reaching 0.59, compared to 0.34 when the reviewers disagreed, as in the latter case WSA predominantly classified deep sleep stages as light sleep. The specificity for light sleep was also starkly improved from 0.56 to 0.72. As a result, the Cohen’s kappa is almost double when reviewers agreed (0.62 vs 0.33 for S/W classification, and 0.52 vs 0.27 for 4-stages classification).

### Sleep Quality With vs. Without PSG

The following analysis aims to explore the impact of PSG on subjective sleep quality. Participants had never done a PSG test before.

Since this analysis does not involve the WSA nor the PSG data, but only the participants’ answers to the questionnaires, it was conducted on the 186 participants for which reported sleep quality data were available. Results are given in Table 6.

**Table 6:** Comparison of Reported Sleep Quality and Sensor Disturbance with and without PSG In both cases, the participant used a WSA and two ScanWatch devices.

|  | **First Night**<br>**With PSG** | **Second night**<br>**Without PSG** |
| --- | --- | --- |
| **Sleep Quality** |  |  |
| **Very poor** | 5.4 % (10) | 1.6 % (3) |
| **Poor** | 34.9 % (65) | 17.2 % (32) |
| **Quite good** | 40.3 % (75) | 38.2 % (71) |
| **Good** | 17.2 % (32) | 31.2 % (58) |
| **Very Good** | 2.2 % (4) | 11.8 % (22) |
| **Sleep Quality compared to usual sleep quality** |  |  |
| **Much worse** | 15.1 % (28) | 3.2 % (6) |
| **Worse** | 37.1 % (69) | 17.2 % (32) |
| **Same** | 41.1 % (77) | 59.7 % (111) |
| **Better** | 5.4 % (10) | 16.7 % (31) |
| **Much better** | 1.1 % (2) | 3.2 % (6) |
| **Sensor Disturbance** |  |  |
| Device that caused sleep disturbances | PSG | WSA |
| **Yes** | 41.4 % (77) | 0 % (0) |
| **No** | 58.6 % (109) | 0 % (0) |

Despite the removal of the nasal cannula and the blood oximeter from our home PSG test, the sleep test was associated with significant discomfort in 41% of the participants during the night of the PSG home test, compared to 0% in the subsequent night with the WSA only. Fifty-two percent (resp. 20%) of the participants reported a degraded sleep quality on the night of the PSG (resp. without the PSG).

Figure 12 illustrates the changes in sleep quality among participants over two consecutive nights. Participants generally experienced better sleep without the PSG sensors, as evidenced by the green transitions going upward from left (first night with PSG) to right (second night without PSG) in the plot. Notable improvements were observed in 52 % of cases, with transitions from fairly good to good (14 %), and from poor to fairly good (16 %), good (6 %), or very good (5 %). Additionally, some participants reported significant improvements from very poor to very good sleep, and others from poor to fairly good. These improvements may be influenced by several factors, including the removal of PSG sensors, which can be intrusive and uncomfortable, potentially disrupting sleep. Furthermore, the awareness of being recorded might alter participants’ sleep behavior, either positively or negatively. The use of unfamiliar devices could also play a role, as participants may initially experience discomfort or anxiety, which diminishes over time. External conditions such as noise and room temperature, which were not controlled in this study, could also significantly impact sleep quality. Therefore, while the data suggests improvements, it is crucial to consider these potential confounding factors when interpreting the results.

**Figure 12:**
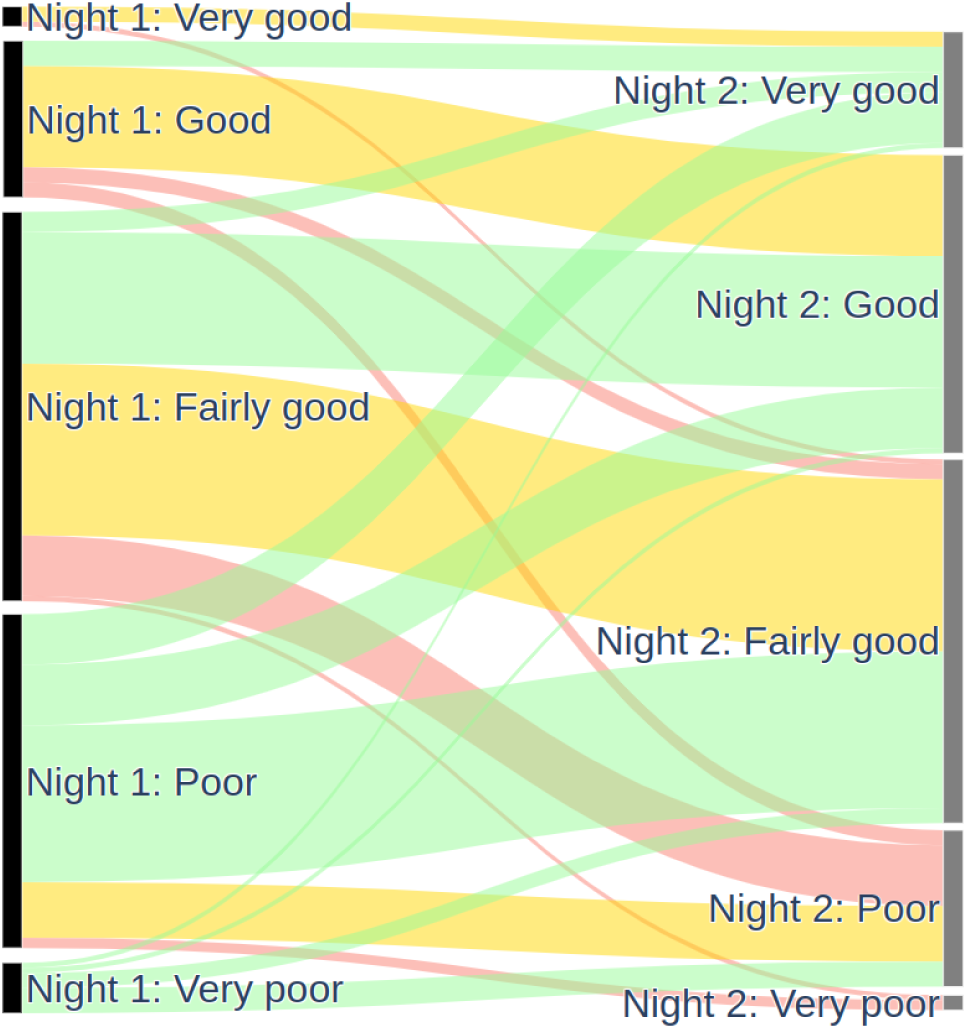
Comparison of Reported sleep quality with and without PSG on 186 subjects. green: improved, yellow: identical, red: deteriorated black: first night with PSG, grey: second night without PSG In both cases, the participant used a WSA and two ScanWatch devices

Table 6 highlights that participants slept as well with WSA as they typically do. On the second night, 59.7% of participants reported the same sleep quality, 16.7% reported better sleep, and 3.2% reported much better sleep compared to their usual experience. This suggests that Withings sleep monitoring devices do not negatively impact sleep quality and may even enhance it for some users.

## Discussion

### Principal Findings

The evaluation of the Withings Sleep Analyzer (WSA) in a home setting for sleep stage detection revealed several key insights. The device demonstrated a high sensitivity of 93% for detecting sleep, underscoring its strong capability to accurately identify sleep periods. However, the specificity was moderate at 73%. The duration of recorded data per individual significantly affects observed performance of wake detection, in a non-trivial way. Excessively short recordings, coinciding with sleep onset and offset, can artificially inflate specificity, consequently increasing accuracy and Cohen’s kappa. This occurs due to a shortening of quiet wake periods hard to distinguish from sleep (reading or screen time in bed, for instance), leading to fewer false positives while true negatives remain relatively constant. Conversely, overly long (e.g., 24-hour) recordings, dominated by easily identifiable diurnal wakefulness, also tend to inflate specificity. In both cases, the performance of the evaluated device is overestimated. The most clinically relevant and challenging scenario for performance evaluation involves analyzing data encompassing the entire in-bed period, including calm wakefulness before sleep onset and after awakening. While AASM guidelines suggest using lights-off and lights-on as proxies for night boundaries, this approach truncates crucial pre- and post-sleep wake data, therefore introducing a bias in TST and classification performance endpoints. To mitigate this, we utilized all available PSG annotations, irrespective of lights-off and lights-on times, to provide a more comprehensive and unbiased assessment of performance across the full in-bed duration.

The classification of sleep stages (into wake, REM, light and deep phases) is more challenging. The confusion matrix Figure 3 shows that 40% of deep sleep phases, 37% of REM sleep, and 20% of wake phases were misclassified as light sleep by the WSA. As a result, the corresponding sensitivities were rather low (respectively 0.56, 0.53, and 0.73). Thirty-two percent of light sleep stages were misclassified, in similar proportions, into REM, deep sleep and wake. In consequence, 1) the REM and deep sleep total durations were underestimated (by circa 30% and 15%), while light sleep duration was overestimated by almost 48% (biases in Table 4), 2) the specificities for wake, REM and deep sleep are very high (0.93, 0.94 and 0.94 respectively), while the specificity for light sleep is moderate (0.69).

We showed that these misclassifications of WSA mirror the disagreements between PSG reviewers, who mainly disagree on classifications between light versus all the other sleep stages (Figure 11). WSA classification was indeed significantly better on epochs where reviewers agreed (79% of the epochs) than epochs where they disagreed. WSA’s sleep stage classification uses an algorithm trained by supervised learning on PSG data annotated by technician experts, so it is expected that WSA performance is limited by the inter-reader variability.

It is customary in assessments of a device performance to also report the accuracy and Cohen’s kappa. We caution that these performance metrics are strongly dependent on the prevalence of the classes (here wake, sleep, REM, deep, light), which creates difficulties in comparing results between studies. In particular for sleep studies, two factors modify significantly the prevalence of the classes. First, as explained above, there is no standardized definition of the beginning of the night, which can significantly modify some performance metrics. Caution must be exercised in comparing studies with significantly different numbers of epochs of wake, as for example in [28,33,38]. Second, the choice of the reference device has some importance, causing more or less sleep disturbances.

With these caveats in mind, the WSA achieved an overall accuracy of 87% for sleep-wake distinction, which is promising for a contactless home monitoring device. Despite this, the accuracy for classifying specific sleep stages was moderate, with REM sleep at 53%, light sleep at 68%, and deep sleep at 56%. The overall accuracy for sleep stage classification was 63%, with a Cohen’s Kappa of 0.49.

Notably, the WSA’s performance was consistent across various demographic subgroups, including different age and BMI categories. Importantly, there was no difference between individuals who slept alone and those with a bed partner, nor between different types or thickness of mattresses. Other groups analyzed included sex, PSQI scores, self-reported sleep quality, and whether individuals woke up during the night, all of which also showed no significant differences in MAE, except for deep sleep proportion by mattress type group. This consistency shows that the device is reliable across diverse patient groups and environments, enhancing its potential utility in a wide range of users.

### Comparison with Prior Work

We compare our results on WSA with influential studies that have contributed to the understanding of consumer sleep-tracking technology. Studies [28], [30] and [34] respectively evaluated seven, five, and six consumer devices against PSG, mainly wearable devices, revealing significant variability in their accuracy for sleep stage detection. [38] compared one contact-free device against PSG. These studies provide a valuable context for evaluating our device’s performance, illustrating both the advancements and challenges in the field of consumer sleep technology. All the data used for this comparison can be found in Supplementary Materials F.

Table 7 summarizes the devices studies, the numbers of participants and numbers of nights, and the performance on all the epochs, using the accuracy and the kappa. When the data was not provided in the study report, we calculated it from the confusion matrix if it was available.

**Table 7:**
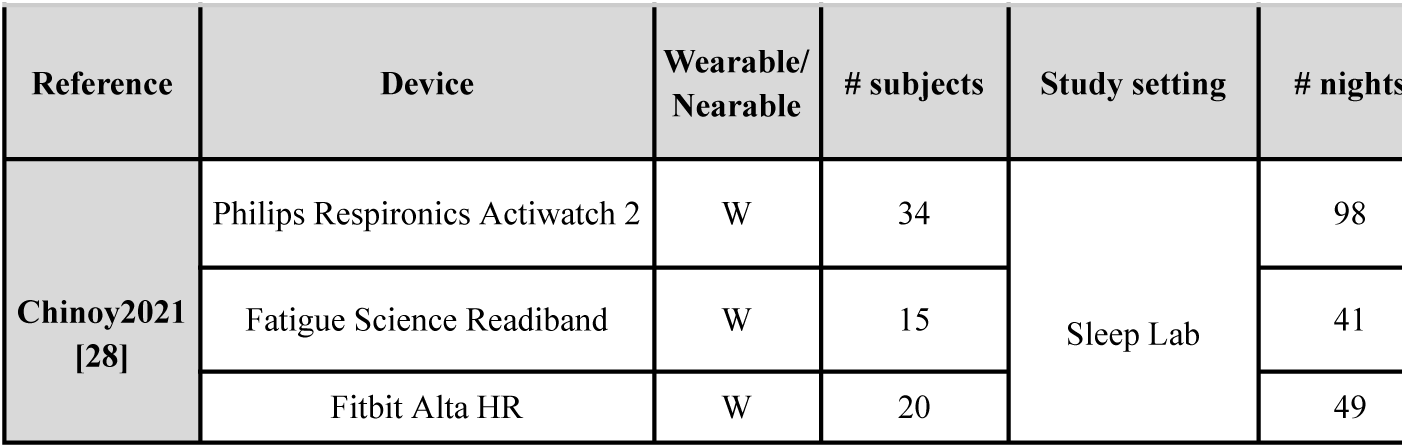
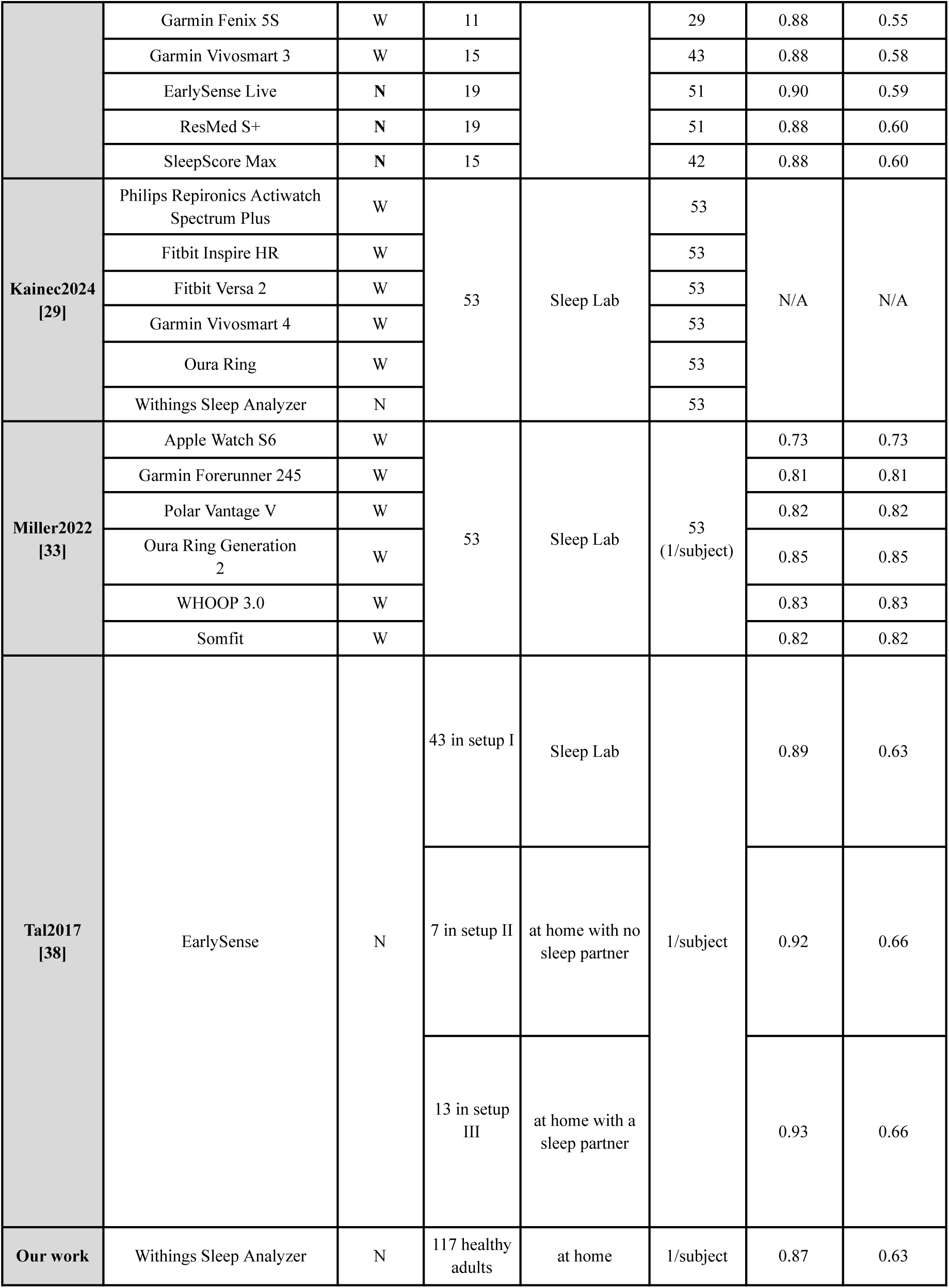
Overview of the Main Devices and Study Parameters in Sleep Monitoring Research N/A: not available S/W : sleep/wake.

We compared the performance metrics reported by other studies on their respective datasets to the performance we obtained on our dataset, without reproducing their results or running their device algorithms on our data.

Our study population is more diverse than in previous research regarding important co-factors of sleep quality, presenting a wider range of age and BMI. In three studies [28,29,33], researchers primarily focused on young adults, only one [38] included a broader age range.

The conditions of our study are not controlled since participants stayed at home, which produced a richer dataset regarding important environmental co-factors of sleep quality, including sleeping arrangements (alone or with a partner), mattress types, ambient temperature, lighting and sound, pre-bedtime activities (eg, no restriction on caffeine or alcohol consumption, on screen time, no forced sleep regularity), activities during time in bed (eg, no forced turning off of the lights). This is reflected, for instance, in a larger range of Total Sleep Time (SD = 71 min) than in other studies (SD in [33 min, 56 min]).

In addition, some studies use strict exclusion criteria also after data collection to ensure data quality and reliability, such as excluding individual nights with a total recording time of less than 7 hours and 50 minutes [28].

Thus, our real-world home setting captures the natural variability in sleep patterns and environmental factors. While this enhances the generalizability of our results, reflecting typical user experiences, it also presents a less favorable scenario for the device, posing additional challenges for accurate sleep stage detection and analysis (for instance activities in bed like reading or screen time can be confounded by the device as sleep). Consequently, our findings provide a more realistic evaluation of the device’s performance in everyday conditions.

#### Sleep-Wake Classification

The sleep sensitivity of WSA is 0.93, similar to most of the other devices whose sensitivity ranges from 0.89 to 0.95 but slightly lower than the Apple Watch S6, Philips Respironics Actiwatch 2, Garmin Fenix 5S, and Vivosmart 3, which achieve sensitivities between 0.98 and 0.99. The WSA’s specificity of 0.73 significantly outperforms most of the other devices, which range from 0.18 to 0.63, although it is slightly lower than the EarlySense device in [0.47, 0.80].

As explained above, the proportion of wake stages in the dataset before and after sleep onset can significantly impact the sleep/wake endpoints. Previous studies vary in the proportions of wake epochs, which refer to the total time spent awake divided by the total recording duration of the entire dataset. Specifically, these proportions ranged from 8 % to 14 % in [28], 15 % in [38], and ranged from 20 % to 33 % in [33], compared to 30 % in our dataset. Lower proportions of wake data can inflate overall accuracies and kappas if sleep epoch classification dominates, as noted in [39]. In datasets with a low proportion of wake epochs, overall accuracy and kappa can be misleadingly high due to the dominance of sleep epoch classification, which are better classified than wake epochs as indicated in the previous paragraph.

With this caveat in mind, let us compare the accuracies and kappas. WSA had an accuracy of 87 % for Sleep/Wake detection. In [28], the EarlySense and ResMed S+ accuracies were 90 % and 88 % respectively [28]. Given the significantly larger proportion of in-bed wake periods in our data, this indicates that the WSA outperforms these devices. Concerning wearable devices, the Fitbit Alta HR achieved 90 % accuracy, the Garmin Fenix 5S, 88 % [28] and the Oura Ring reached 83 % accuracy [33]. Again, given that our data have the largest proportion of wake epochs, these results indicate the superior accuracy of WSA.

WSA’s sleep-wake Cohen’s Kappa value is 0.63, indicating substantial agreement and outperforming ranges reported in [33] (0.27 to 0.56) and [28] (0.25 to 0.51). This value is close to the EarlySense device from [38], which has a Kappa of 0.68.

#### Sleep Stages Classification

The light sleep sensitivity of WSA is 0.68, just as Garmin Fenix 5S, Garmin Forerunner 245, SleepScore Max, ResMed S+, Whoop 3.0, and Somfit, therefore outperforming Apple Watch, Polar Vantage V, Oura Ring, and EarlySense which achieve sensitivities of 0.44, 0.60, 0.66 and in [0.57, 0.68], although it is lower than Garmin Vivosmart 3 and Fitbit Alta HR, which achieve light sleep sensitivities of 0.70 and 0.76.

Most of the devices have significantly lower deep sleep sensitivities than light sleep’s, ranging from 0.33 to 0.62 for Garmin Forerunner 245 (0.28), Polar Vantage V (0.33), Fitbit Alta HR (0.53), WSA (0.53), Garmin Fenix 5S (0.56), Garmin Vivosmart 3 (0.56), ResMed S+ (0.59), SleepScore Max (0.59), Oura Ring Gen 2 (0.62). The exceptions are EarlySense and Whoop 3.0 devices that show similar deep sleep sensitivities to light sleep’s, respectively in [0.52, 0.68] and of 0.62.

Regarding REM sleep, the WSA shows a sensitivity of 0.54, similar to Garmin Vivosmart 3 (0.54) and EarlySense devices ([0.4, 0.64]). It therefore outperforms SleepScore Max and Polar Vantage V, both at 0.49, Garmin Fenix 5S, ResMed S+, Garmin Forerunner 245 all three having a REM sleep sensitivity of 0.50 and Oura Ring that reaches a sensitivity of 0.52. However, these devices are outperformed by Somfit, Whoop 3.0 and Fitbit Alta HR that achieve sensitivities of 0.60, 0.66 ad 0.69.

The Apple Watch cannot distinguish deep and REM sleep and achieves a (deep sleep and REM) sensitivity of 0.71.

A better sensitivity in a given sleep stage compared to the other devices is often counterbalanced by a worse sensitivity in another stage; for instance, Whoop device high REM sleep sensitivity of 0.66 faces a low 0.58 light sleep sensitivity, just like Garmin Forerunner 245 high light sensitivity of 0.68 is offset by a low deep sleep sensitivity of 0.28. Still, other devices stand out from the others with balanced performance across light, deep and REM sleep sensitivities: Fitbit Alta HR, Oura, Somfit, Garmin Vivosmart 3, ResMed S+, SleepScore Max, and WSA.

The WSA, with an accuracy of 0.63 for sleep stage classification, outperforms most of the other devices whose accuracies are in the range [0.43, 0.60]. The WSA has a similar accuracy to EarlySense ([0.59, 0.66]) and has a lower accuracy than the Fitbit Alta HR and Somfit with 0.68 and 0.64 accuracies. The WSA’s kappa of 0.50 is one of the highest, similar to Somfit (0.50), EarlySense ([0.40, 0.50]), and Fitbit Alta HR (0.51), and below Whoop (0.57), therefore outperforming the other devices which kappas range in [0.20, 0.46].

The confusion matrices show that, for all devices, the highest proportion of misclassification for each stage is wrong light sleep detection, which can be explained by the inherent difficulty in distinguishing light from the other sleep stages by human experts, as shown in the Results section.

#### Device Comparison Conclusion

When comparing sleep monitoring devices, while absolute results may not appear exceptionally high, the performance of the WSA surpasses the other devices in the market in sleep-wake distinction. Regarding sleep stage classification, the WSA is on par with similar products. This is noteworthy given the challenges in accurately classifying sleep stages, complicated by reviewer disagreements and limitations of the current gold standard, making it a complex issue. Despite these challenges, the WSA demonstrates competitive performance, highlighting its reliability in a field often debated for the accuracy and consistency of its gold standard. As detailed before, a given device’s strength is the weakness of another device. The selection of the sleep monitoring device must be guided by the researcher or clinician’s specific goals.

### Methodological Aspects and Strengths of the Study

The rapid adoption of new sleep technologies in consumer and clinical settings demands rigorous performance evaluation [40]. To improve the reliability and comparability of sleep research, it is essential to draw on existing publications that offer guidance on harmonizing methods and implementing a uniform analysis pipeline across different studies [31–33]. Our study was designed to resolve methodological issues frequently found in the literature.

The study’s strength lies first in its diverse sample of over 100 participants, who are for the most part healthy and with diverse demographic characteristics. Additionally, the inclusion of relevant factors that may negatively impact the device’s performance, like the presence of a bed partner and variations in mattress type and thickness, further enriches the dataset, allowing for a comprehensive evaluation of the device’s robustness across different real-life conditions in a healthy population. This allowed us to conduct meaningful sub-group analyses.

A second strength of the study is the method of PSG analysis. Unlike [33] which opted for a single reviewer, or two reviewers without consensus as in [28], our study is aligned with [29] by implementing a double review process with adjudication by a third reviewer in case of disagreement. This consensus methodology is preferable since it enhances the reliability of the sleep stage classification. In particular, we have shown that reviewers often err by misclassifying light sleep stages, a finding consistent with previous research reporting that the inter-rater reliabilities for light and deep stages were moderate [39]. A reliable assessment of devices’ performance in sleep stage classification therefore requires a strong methodology to establish the ground truth.

Third, with a view to limit the participants’ discomfort, we chose to perform PSG at home with a limited number of sensors, without the nasal cannula and oximeter. This setup reduces the sleep disturbances typically associated with lab-based PSG, allowing for a more natural assessment of sleep patterns. This limited set of sensors is sufficient for the evaluation of TST and sleep stage classification, with the trade-off that sleep onset, time in bed and wakefulness after sleep onset cannot be assessed. Yet, significant sleep disturbances were reported by the participant during the PSG test, underscoring the challenges of completely undisturbed monitoring even in home environments. Despite this, the study offers a realistic evaluation of the WSA’s capabilities under typical sleeping conditions

Fourth, we aimed to give a comprehensive report of the device’s performance. We therefore designed the study to allow for epoch-by-epoch data collection, which is crucial for detailed temporal analysis since it enables a granular examination of sleep patterns and the consistency of measurement errors over time. This level of detail is necessary to accurately assess the reliability and utility of sleep monitoring devices in real-world settings. Furthermore, we repeatedly emphasized that it is dangerous to summarize a performance with one particular performance metric, and that the composition of the dataset should be factored in when performing comparisons between studies. We explained how biases can easily be introduced in such studies. Thus, transparency requires a presentation of the results including all the performance endpoints (sensitivity, specificity, accuracy and Cohen’s kappa, mean absolute error and Bland-Altman analyses), each presenting a different facet of the device’s performance.

In conclusion, given these methodological strengths, we think our study provides valuable insights into the WSA sleep stage algorithm’s performance in real-world settings.

### Challenges and Limitations

Despite our efforts for a solid methodology, the study faced several challenges.

#### Challenges in Defining Bedtime and Wake Time in Sleep Studies

A first challenge is created by the lack of a clear-cut, or standardized, definition of the start time and end time of a sleep study. As per AASM guidelines [35], bedtime and wake time are defined as the “lights-off” and “lights-on” respectively. This is not adapted to studies in real-world settings, where subjects often go to bed and engage in activities before attempting to fall asleep. In everyday life, individuals may not turn off the lights immediately upon going to bed, nor do they always intend to sleep right away. Similarly, in the morning, people may not turn on the lights immediately upon awakening, and their actual wake time may not coincide with the “lights-on” reference. This discrepancy is compounded by the lack of position sensor, which makes it difficult to accurately determine when a person *intends* to sleep. In other studies [28], start and end times of sleep are determined by the subjects themselves, as suggested in standard sleep diaries [41], but the precision of this method is poor.

This lack of standardization is detrimental to the assessment of nearable and wearable devices intended to be used in home settings. Errors in sleep estimation, where wakefulness is mistakenly classified as sleep, are more frequent at the beginning and end of the night. The reason is that many devices, including the WSA, rely heavily on movement detection to distinguish wake from sleep. Thus wake periods when the subject is immobile and calm in bed, such as when reading or scrolling with a low heart rate, are frequently mistaken as sleep by the device.

Additionally, the practice of activating the PSG immediately before sleep and deactivating it early prevents from gathering comprehensive data on a subject’s sleep habits. This includes behaviors such as spending extended periods in bed without the intention to sleep. To address this limitation, future studies could extend the duration of PSG recordings to capture activities that occur between the time a person gets into bed and falls asleep, as well as between waking and getting out of bed. This approach would provide a more complete picture of sleep behaviors and enable a thorough evaluation of sleep stage classification algorithms in real-world settings.

#### Challenges in Data Collection from a Nearable

A second challenge in studies with connected devices is data loss. Our study is no exception since nearly 19% of the data were lost, primarily due to the wireless transfer by WSA of a large amount of raw sensor data (250 samples per second). Note that this high rate is not representative of the intended use of the device, which in its commercial version only transfers processed data at a low frequency. To mitigate the risk of data loss, Wi-Fi routers paired with the WSA were installed at the subject’s home. The routers experienced occasional connectivity problems. Data loss was further exacerbated by issues such as accidental unplugging of the device and improper reconnection without calibration. These technical challenges underscore the difficulties of gathering high-quality, high-frequency data in uncontrolled settings.

Another factor impacted the accuracy of sleep stage classification: despite receiving instructions from a technician, participants with large beds did not always sleep above the sensor but on the other side of the bed. In such cases, the WSA is not operated optimally, which may have restricted its performance.

### Future Work

The promise of nearable and wearable devices like the WSA is to facilitate the longitudinal monitoring of subjects in their home environment. While the results of this study are conclusive, it remains a challenge to demonstrate the added value of such devices for longitudinal follow-up of patients when the gold standard itself has serious limitations. To validate the utility of longitudinal passive data collection, it is essential to demonstrate that errors in sleep quality measures are consistent over time for a given user. This means that the bias observed in these measures should remain stable across different time points for the same individual, ensuring that the device is both useful and accurate in analyzing temporal data. However, this assessment would require participants to sleep with PSG equipment over multiple nights, which is not only costly but also disruptive to natural sleep patterns. This highlights the need for innovative methodologies that can reliably assess and ensure the consistency of bias in sleep quality measures over time without the drawbacks of traditional PSG setups. In addition, our study population only includes healthy participants. There is no data on the performance of the device in special populations that might benefit the most from a longitudinal follow-up.

## Conclusion

In this study, we aimed to 1) assess the performance of a contactless device for sleep stage identification against gold standard PSG, 2) benchmark it against other comparable devices, and 3) better understand the limitations of such devices. We also hope to have made a strong point about important methodological issues.

The WSA is a valuable and easy-to-use tool with sleep stage identification performance comparable to similar products on the market and outperforming them for sleep-wake distinction. Being contactless, the primary interest of the WSA lies in the monitoring of temporal evolution of various metrics, such as heart rate, respiratory rate, heart rate variability, nocturnal activity, apnea-hypopnea index, and snoring. In particular, the WSA proved itself to be useful in longitudinal studies to explore associations with conditions such as apnea-hypopnea index (AHI), insomnia, and correlations with hypertension [13,40,42,43].

Looking ahead, the WSA holds significant potential for a wide range of research and clinical applications. It can be instrumental in investigating the impact of sleep quality and physical activity on blood pressure variability, analyzing large-scale in-home sleep data to assess sleep irregularity and its association with hypertension [44,45], and examining sleep characteristics in relation to night-time voids in non-patient populations [46]. The WSA’s capability to monitor sleep in natural environments [47] underscores its versatility and relevance in advancing our understanding of sleep health and its implications.

## Data Availability

The data that support the findings of this study were collected during a clinical study (Approval Reference Number: 2018-A03129-46, French Institutional Review Board) conducted by Withings. These data are proprietary and due to their sensitive nature as human clinical data, they are not publicly available.

## Acknowledgements

We would like to acknowledge Paul Edouard and Sébastien Ducrot for their crucial support and technical expertise in the offline run of the algorithm.

## Acronyms

AASM: American Academy of Sleep Medicine
BMI: Body Mass Index
CE: European Conformity (Conformité Européenne)
ECG: Electrocardiogram
EEG: Electroencephalogram
EOG: Electrooculogram
EMG: Electromyogram
FDA: Food and Drug Administration
MAE: Mean Absolute Error
NEDeep: Number of Episodes in Deep sleep stage
NELight: Number of Episodes in Light sleep stage
NEREM: Number of Episodes in REM sleep stage
OLS: Ordinary least squares
PIDeep: Proportion of the night in Deep sleep stage
PILight: Proportion of the night in Light sleep stage
PIREM: Proportion of the night in REM sleep stage
PSG: Polysomnography
PSQI: Pittsburgh Sleep Quality Index
RANSAC: Random sample consensus algorithm
REM: Rapid Eye Movement (sleep stage)
RPSGTs: Board of Registered Polysomnographic Technologists
TIDeep: Time spent in Deep sleep stage
TILight: Time spent in Light sleep stage
TIREM: Time spent in REM sleep stage
TST: Total Sleep Time
WSA: Withings Sleep Analyzer

