## Supplementary Materials A to D for "Evaluation of a Contactless Sleep Monitoring Device for Sleep Stage Detection against Home Polysomnography in a Healthy Population"

### Table of Contents

**Table of Contents** **1**
**Supplementary Materials A — Methods** **2**
Data Processing 2
Reference 2
Smoothed, PSG-based Consensus 2
Synchronization 3
Definitions: Start and End of the Night 3
**Supplementary Materials B — Performance Evaluation Metrics and Tools** **5**
Mean Absolute Error on Sleep Metrics 5
The mean absolute error (MAE) quantifies the accuracy of a model or prediction by calculating the average of the absolute differences between predicted values and actual values. 5
Classification Metrics 5
Plots 6
Regression 6
Bland-Altman Plots 7
**Supplementary Materials C — Subgroup Analyses** **8**
**Supplementary Materials D — Results** **9**
**References** **19**

### Supplementary Materials A — Methods

#### Data Processing

##### Reference

Each PSG was read by two trained reviewers following AASM scoring rules [1]. To limit the impact of inter-reader differences [2], epochs with disagreements were adjudicated by a third qualified professional. The adjudicated review is henceforth called the “reference” or the “consensus”.

##### Smoothed, PSG-based Consensus

The raw, PSG-based consensus data consists of manually labeled 30-second epochs. WSA outputs a new sleep stage every minute. By design, short stages are smoothed out over 6 minute windows. Therefore, to make a meaningful comparison of the hypnograms produced by WSA and the reference method, we 1) upsampled the output of WSA sleep stage detection algorithm to 30-second epochs by repetition of the value, 2) processed consensus data to get a smoothed version of the PSG-based consensus (see below), and 3) compared the upsampled output of the WSA sleep stage classification algorithm to the smoothed version of the PSG-based consensus. The smoothing process of the PSG-based consensus consists in determining the majority sleep stage over a 6-minute window centered around each 30-second epoch. This approach effectively reduces the variability of the PSG hypnogram by ignoring non-majority epochs within each 6-minute phase, as illustrated on Figure A.XX. With this method, all events of one sleep stage lasting less than 1min30s and most of those lasting less than 2min30s are removed.

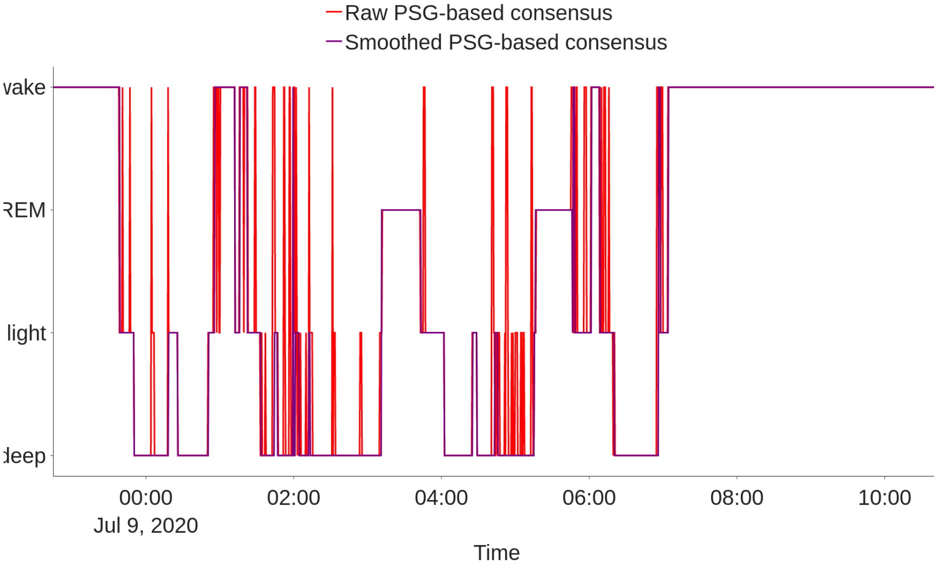


*Figure A.1: Illustration of consensus hypnogram smoothing (subject 20200708_1)*

#### Synchronization

Recordings from the PSG and the WSA are synchronized using the respiratory signals. The WSA respiratory signal was computed from the normalized pressure signal bandpass-filtered in the frequency range [0.19 Hz, 1 Hz], and the respiratory signal of the PSG was obtained from the ECG filtered similarly [2]. Synchronization then consisted in identifying the optimal affine time warping that maximized their correlation, followed by a visual inspection to ensure that the correlation had a sharp extremum (compare Figures A.2 and A.3, only the former presents an extremum, for a “shift value” around -5000).

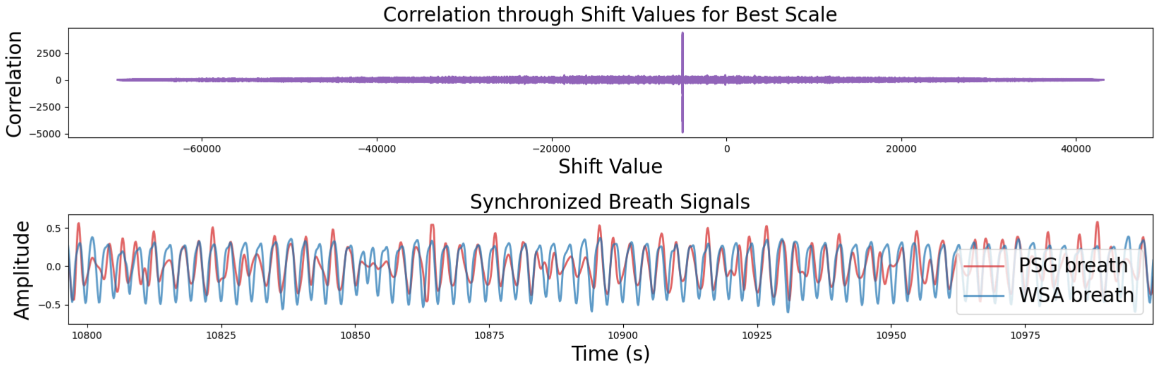


*Figure A.2: Illustration of a valid synchronization (subject 20201118_1)*

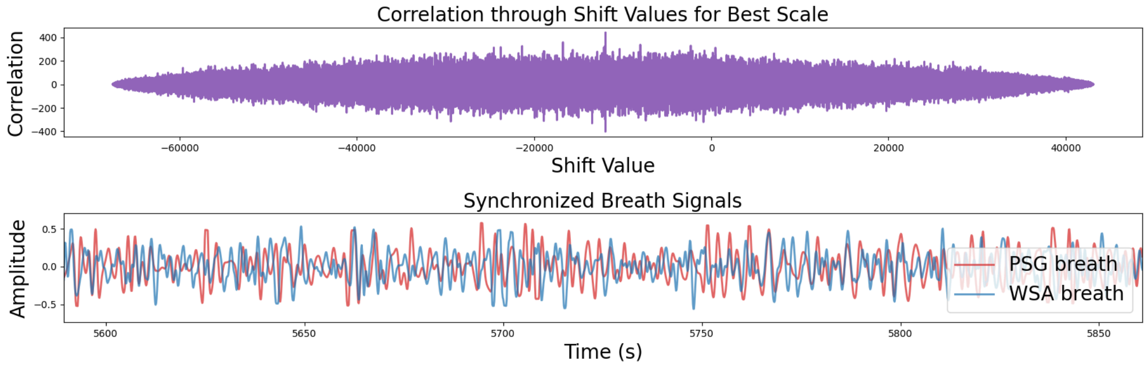


*Figure A.3: Illustration of an invalid synchronization (subject 20200916_1)*

Once determined, these coefficients were applied to the WSA hypnogram data to synchronize it with the reference consensus data.

#### Definitions: Start and End of the Night

The polysomnograph used in this study recorded 3 EEG, 1 ECG, 2 EOG, 1 submental EMG, and ambient light. In particular, it did not have a position sensor, so we do not have a reference for time in bed and sleep related measures depending on it (bedtime, sleep latency, wakefulness after sleep onset, sleep efficiency, etc.).

In the absence of an indicator of the subject's presence in bed in PSG data, lights-off/on events were used as a proxy to estimate bed presence. If participants went to bed before starting the PSG, or got up after stopping the PSG, the WSA might detect presence without corresponding PSG data. Such unmatched data were excluded from the analysis, relying solely on WSA data that could be compared to the reference.

### Supplementary Materials B — Performance Evaluation Metrics and Tools

#### Mean Absolute Error on Sleep Metrics

The mean absolute error (MAE) quantifies the accuracy of a model or prediction by calculating the average of the absolute differences between predicted values and actual values.

#### Classification Metrics

In evaluating the performance of the sleep staging algorithm, we employ standard classification metrics. These metrics, computed with the ground truth obtained from the PSG and predictions from the WSA, provide insights into the algorithm's ability to correctly classify sleep stages. Below are the definitions and descriptions of each metrics.

**Confusion Matrix Example**
To illustrate these metrics, consider a binary classification scenario (e.g., sleep vs. wake) as in Figure B.1, using the following acronyms:

- True Positives (TP): Instances correctly identified as wake
- False Positives (FP): Instances incorrectly identified as wake
- True Negatives (TN): Instances correctly identified as sleep
- False Negatives (FN): Instances incorrectly identified as sleep

| | | **Ground Truth (PSG)** | |
|---|---|---|---|
| | | Sleep | Wake |
| **Predictions (WSA)** | Sleep | TN | FN |
| | Wake | FP | TP |

*Table B.1: Confusion matrix of a sleep-wake classification*

This confusion matrix serves as a valuable tool for evaluating the performance of the classification model, providing a clear visualization of the algorithm's strengths and weaknesses in identifying sleep and wake states.

Sensitivity (Recall) $S_e$: The proportion of actual positive cases (eg, awake time) correctly identified by the Device Under Test (DUT). It is calculated as:

$$S_e = \frac{\text{TP}}{\text{TP} + \text{FN}}$$

Specificity $S_p$: The proportion of actual negative cases (eg, sleep time) correctly identified by the DUT. It is calculated as:

$$\mathrm{Sp} = \frac{\mathrm{TN}}{\mathrm{TN} + \mathrm{FP}}$$

Accuracy: The proportion of all cases (both positive and negative) that are correctly identified. It is calculated as:

$$\mathrm{Acc} = \frac{\mathrm{TP} + \mathrm{TN}}{\mathrm{Total}}$$

Kappa: A statistical measure of inter-rater agreement for qualitative (categorical) items, accounting for agreement occurring by chance.

$$\kappa = \frac{P_o - P_e}{1 - P_e}$$

where:

$P_o$ is the observed agreement, which is the proportion of instances where the raters agree (ie, the sum of the diagonal elements of the confusion matrix divided by the total number of instances).

$P_e$ is the expected agreement by chance, calculated based on the marginal totals of the confusion matrix.

These metrics were calculated for each night individually and then averaged across all nights. This method captures night-to-night variability, providing a nuanced assessment. It allows for detailed analysis of specific nights, offering granular insights into patterns or issues.

#### Plots

##### Regression

Ordinary Least Squares (OLS) regression was used to model the relationship between estimated sleep metrics and reference measurements. The data were assessed for normality and homoscedasticity of the residuals were assessed. Normality was evaluated using the Shapiro-Wilk test, while homoscedasticity was assessed using Levene's test. Both tests confirmed that the assumptions of normality and homoscedasticity were met for all metrics except total sleep time (TST), allowing for the application of parametric statistical methods in subsequent analyses. Regarding TST, RANSAC (Random sample consensus) regression has been performed as it is robust to outliers and does not assume constant error variance, making it suitable for data with varying error distributions [5].

For each regression, the following values are provided:

- R-squared value: “proportion of variance in the outcome variable which is explained by the predictor variables in the sample” [3]
- p-values of the slope and the intercept: “probability under the assumption of no effect or no difference (null hypothesis), of obtaining a result equal to or more extreme than what was actually observed” [4]

#### Bland-Altman Plots

Bland-Altman plots are used to assess the agreement between two different measurement techniques. They display the difference between the measurements on the y-axis against their mean on the x-axis. Bland-Altman plots are an effective method for comparing estimated sleep metrics to reference measurements by visually assessing the agreement between the two. They highlight the mean difference and the limits of agreement, revealing any systematic bias or variability.

### Supplementary Materials C — Subgroup Analyses

| Metadata name | Meaning | Categories |
|---|---|---|
| age_group | Age category the participant belongs | 18-30, 30-50, 50+ |
| bmi_group | BMI category the participant belongs | <18.5, 18.5-25, 25-30, >30 |
| sex | Participant's sex | man, woman |
| is_couple | Did the participant have a bed partner the night of the recording? | no, yes |
| psqi_score_group | Results obtained using the Pittsburgh Sleep Quality Index (PSQI) survey, categorized into 4 groups | no_sleep_difficulty, mild_sleep_difficulty, moderate_sleep_difficulty, severe_sleep_difficulty |
| psqi_quality_group | PSQI Component 1: Subjective sleep quality | good, bad |
| psqi_latency_group | PSQI Component 2: Sleep latency | good, bad |
| psqi_duration_group | PSQI Component 3: Sleep duration | good, bad |
| psqi_efficiency_group | PSQI Component 4: Habitual sleep efficiency | good, bad |
| psqi_disturbances_group | PSQI Component 5: Sleep disturbances | good, bad |
| psqi_medication_group | PSQI Component 6: Use of sleeping medications | good, bad |
| psqi_day_dysfunction_group | PSQI Component 7: Daytime dysfunction | good, bad |
| hypnotics | Did the participant take hypnotics the night of the recording? | no, yes |
| sleep_quality | Self-reported sleep quality the night of the recording | very good, good, quite good, bad, very bad |
| mattress | Type of the mattress the participant slept on the night of the recording | spring, memory foam, foam, latex, other, unknown |
| mattress_thickness_group | Thickness of the mattress the participant slept on the night of the recording | thin (<20 cm), thick (≥20 cm) |
| had_arousals | Did the participant wake up at least once during the night of the recording (self-report)? | no, yes |

*Table C.1: Summary of subgroup categories used in boxplots obtained on standard sleep-related metrics*

### Supplementary Materials D — Results

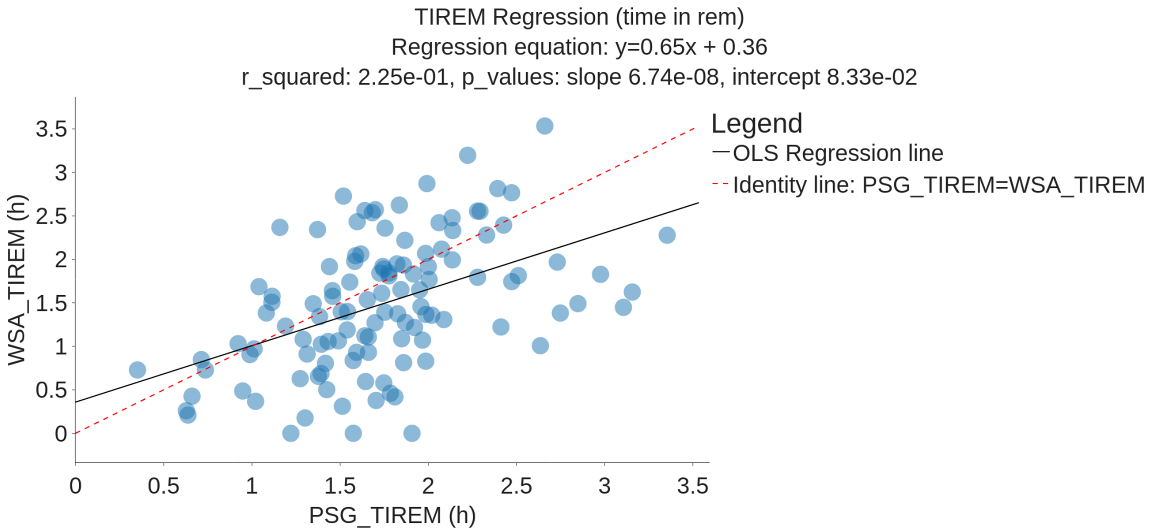


*Figure D.1: Time In REM (TIREM): OLS Regression plot between WSA and PSG (black line) compared to the identity line (red dashed line) on 117 subjects ($r^2 = 0.65$, slope p-value: 7e-8)*

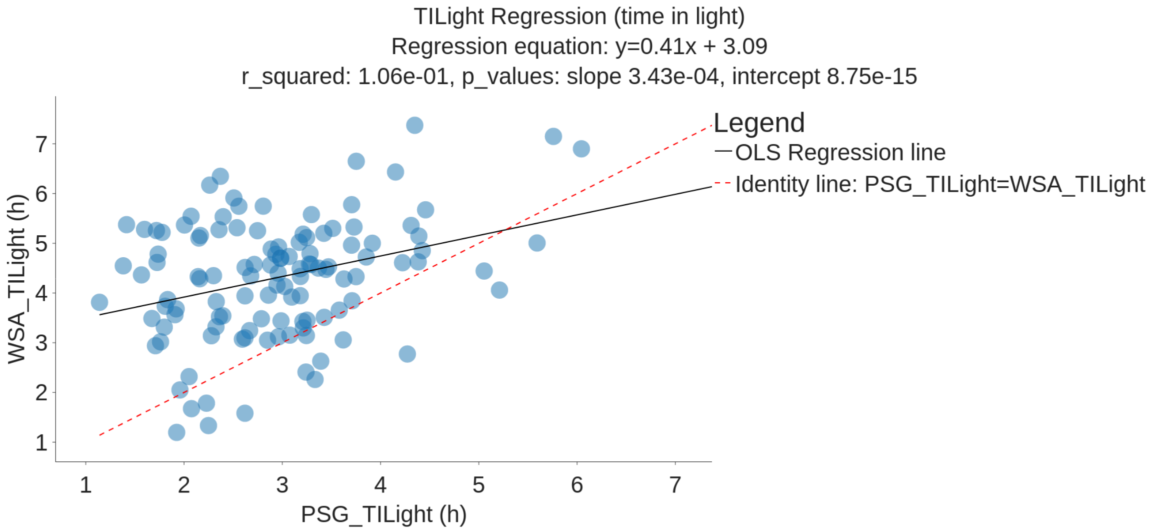


*Figure D.2: Time In Light sleep (TILight): OLS Regression plot between WSA and PSG (black line) compared to identity line (red dashed line) on 117 subjects ($r^2 = 0.41$, slope p-value: 3e-4)*

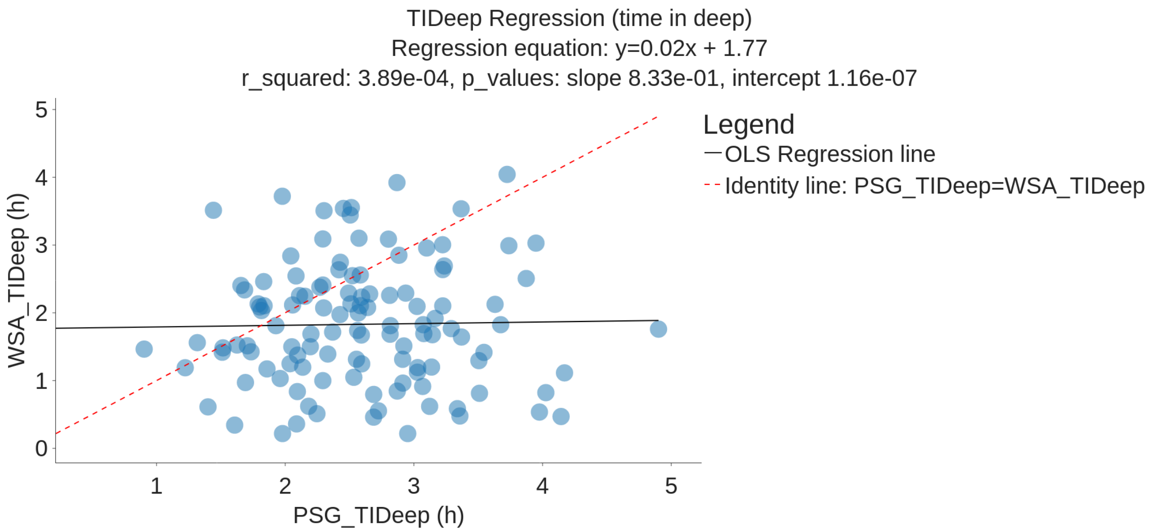


*Figure D.3: Time In Deep sleep (TIDeep): OLS Regression plot between WSA and PSG (black line) compared to identity line (red dashed line) on 117 subjects ($r^2$ = 0.02, slope p-value: 8e-1)*

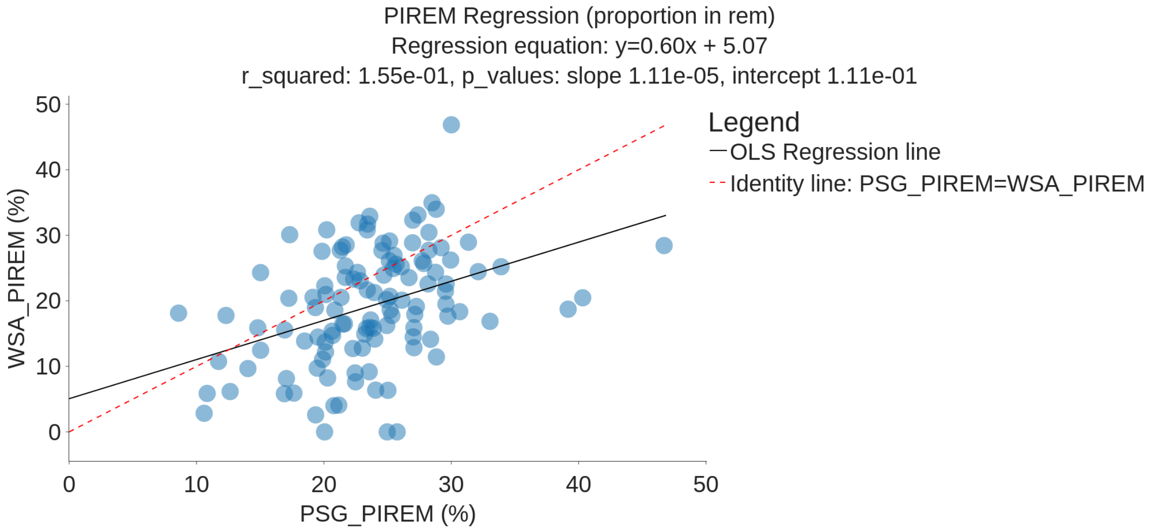


*Figure D.4: Proportion In REM (PIREM): OLS Regression plot between WSA and PSG (black line) compared to identity line (red dashed line) on 117 subjects ($r^2$ = 0.60, slope p-value: 1e-5)*

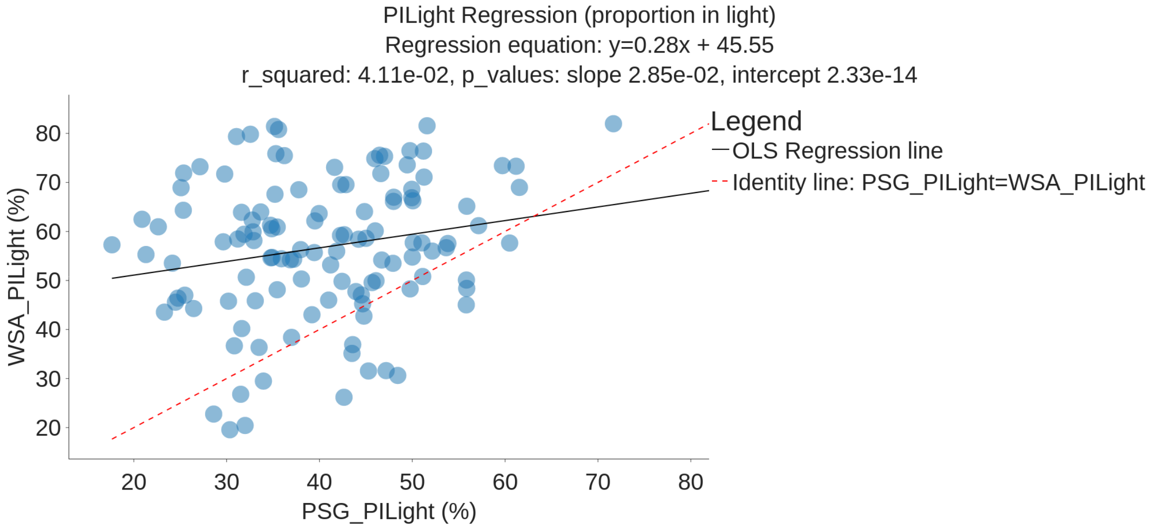


*Figure D.5: Proportion In Light sleep: OLS Regression plot between WSA and PSG (black line) compared to identity line (red dashed line) on 117 subjects ($r^2$ = 0.28, slope p-value: 3e-2)*

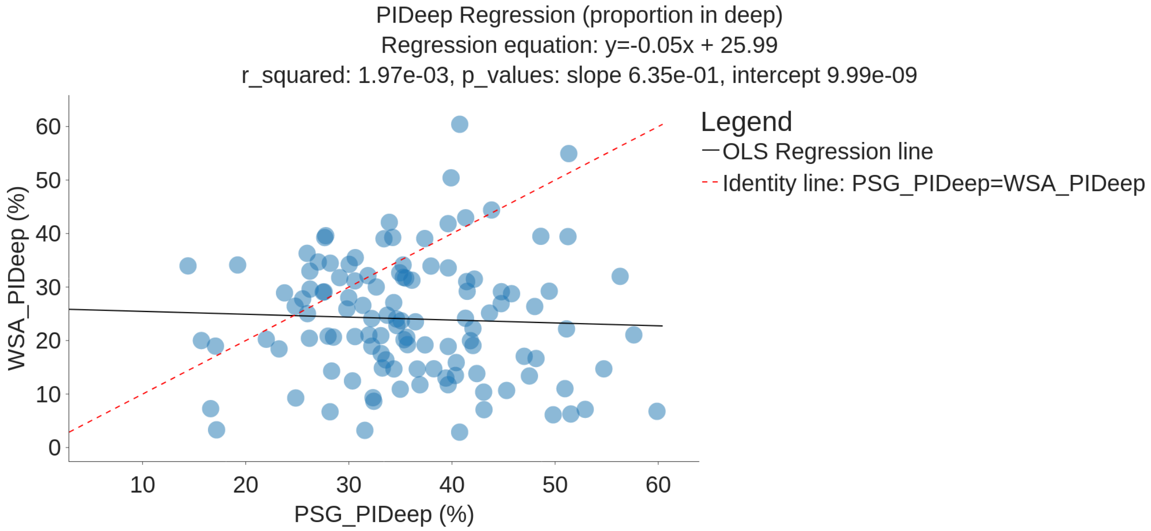


*Figure D.6: Proportion In Deep sleep (PIDeep): OLS Regression plot between WSA and PSG (black line) compared to identity line (red dashed line) on 117 subjects ($r^2$ = 0.05, slope p-value: 6e-1)*

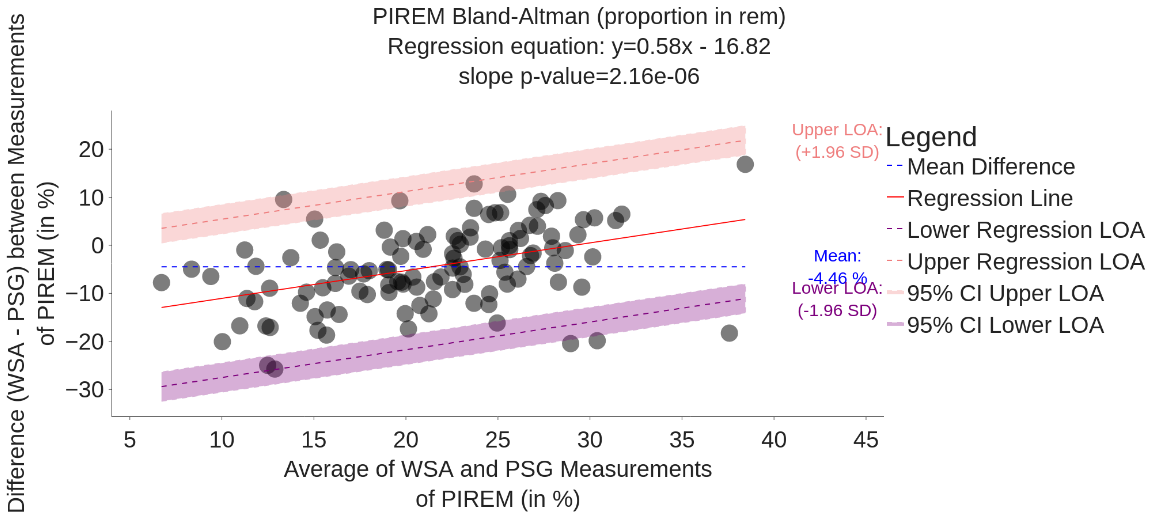


*Figure D.7: Bland-Altman Plot Comparing WSA and PSG Estimation of PIREM (%) on 117 subjects*

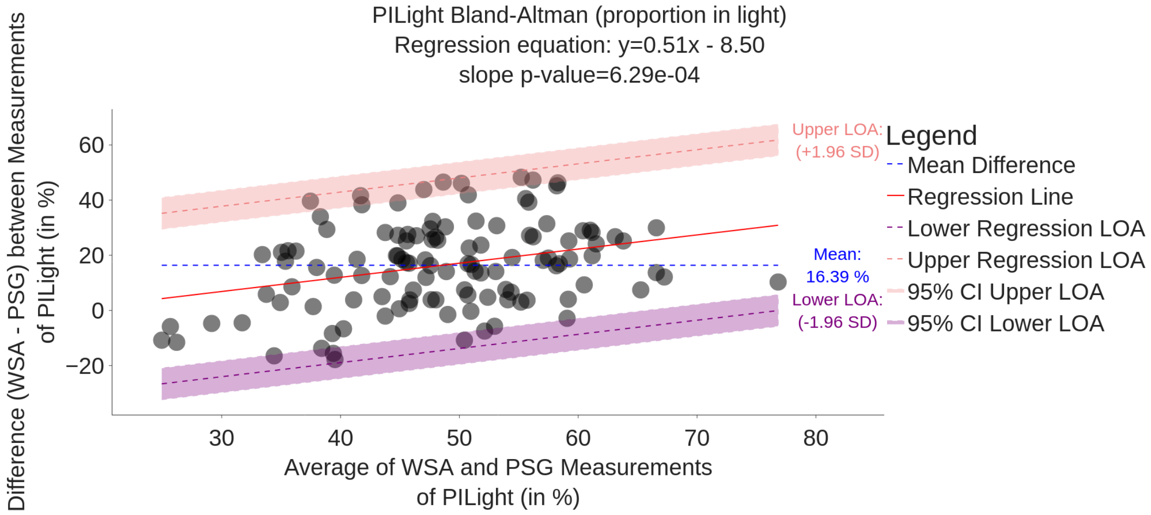


*Figure D.8: Bland-Altman Plot Comparing WSA and PSG Estimation of PILight (%) on 117 subjects*

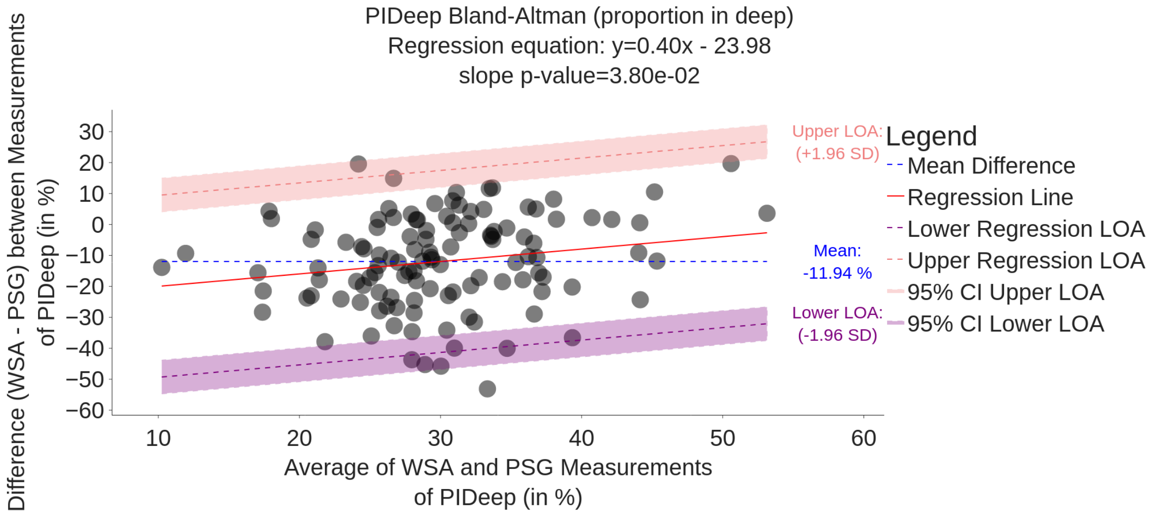


*Figure D.9: Bland-Altman Plot Comparing WSA and PSG Estimation of PIDeep (%) on 117 subjects*

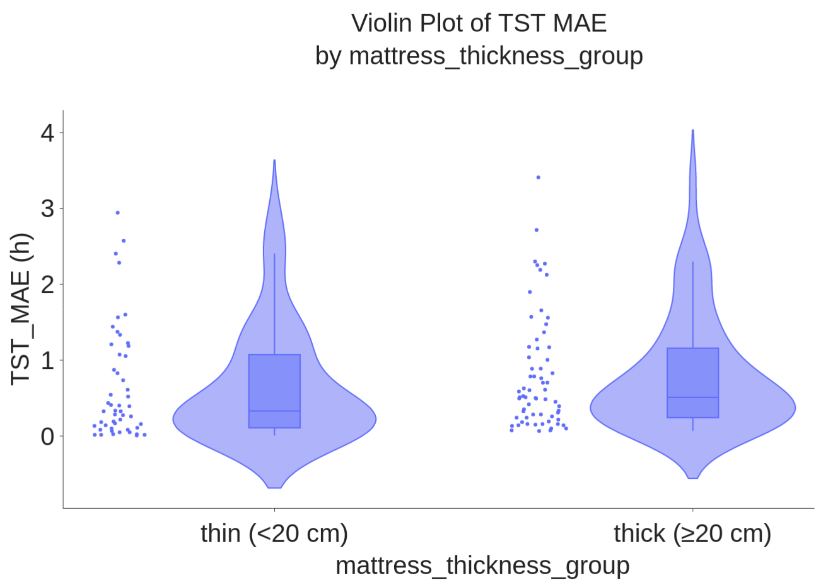


*Figure D.10: Boxplot of TST MAE (h) by mattress thickness group on 117 subjects*

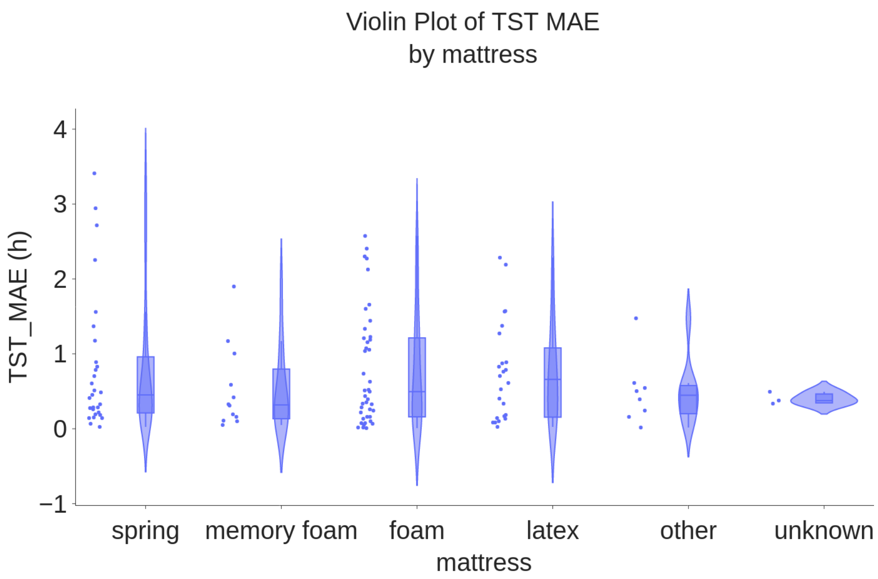

*Figure D.11: Boxplot of TST MAE (h) by mattress type group on 117 subjects*

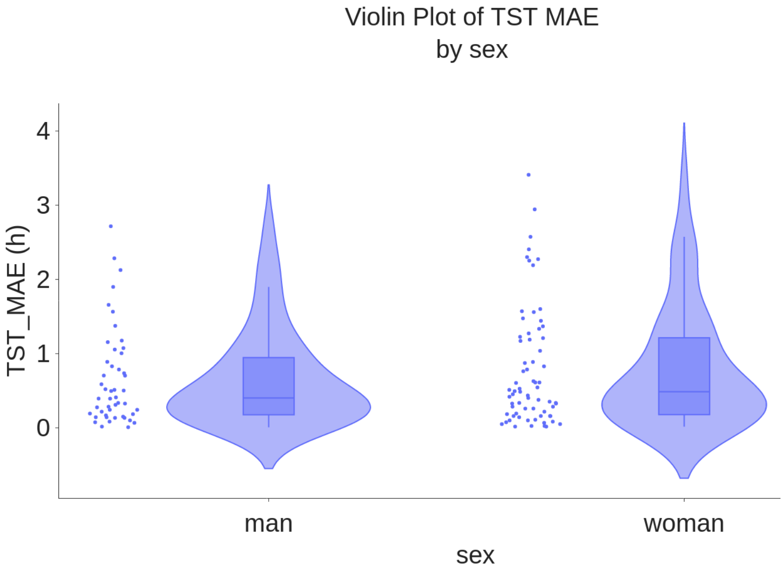


*Figure D.12: Boxplot of TST MAE (h) by sex group on 117 subjects*

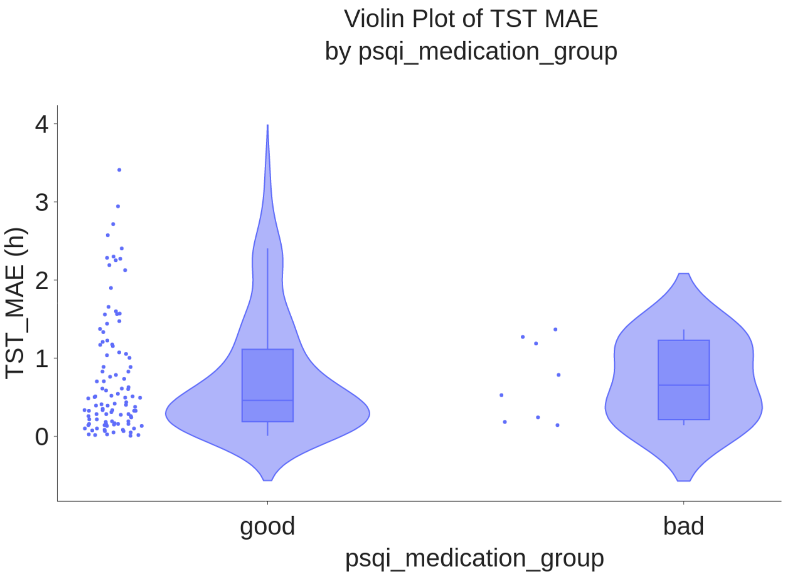

*Figure D.13: Boxplot of TST MAE (h) by PSQI medication group on 117 subjects*

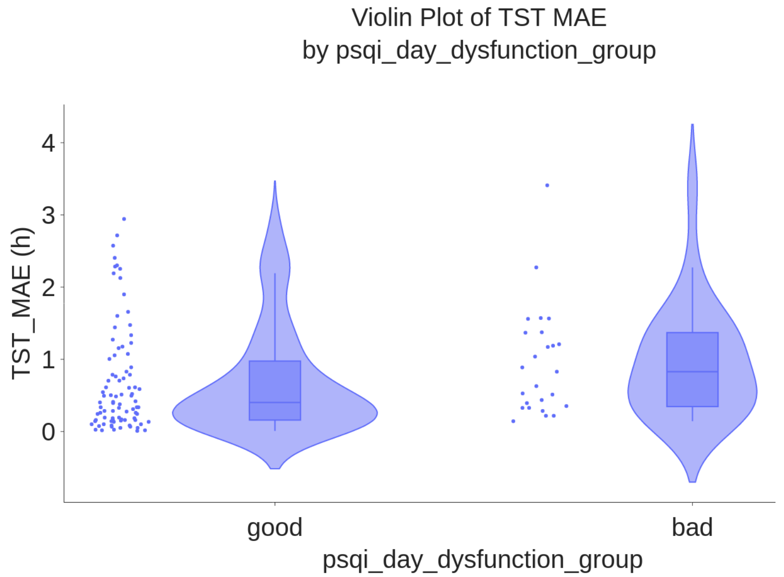


*Figure D.14: Boxplot of TST MAE (h) by PSQI day dysfunction group on 117 subjects*

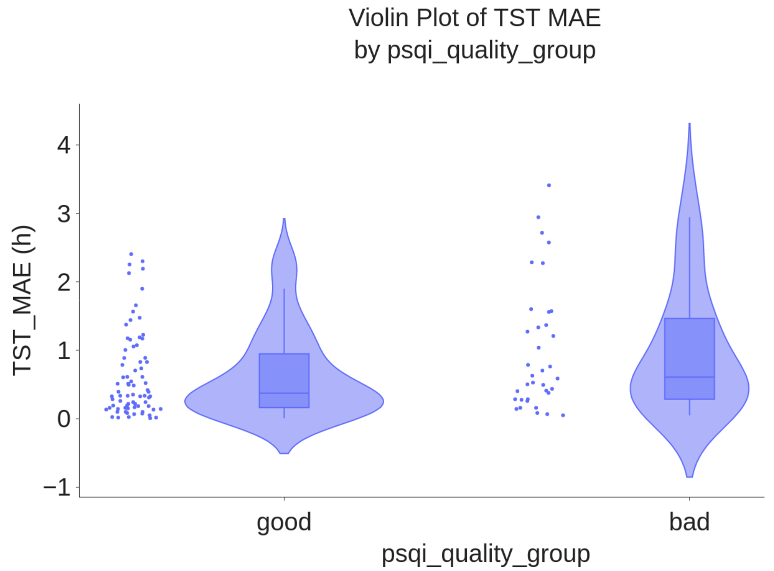


*Figure D.15: Boxplot of TST MAE (h) by PSQI sleep quality group on 117 subjects*

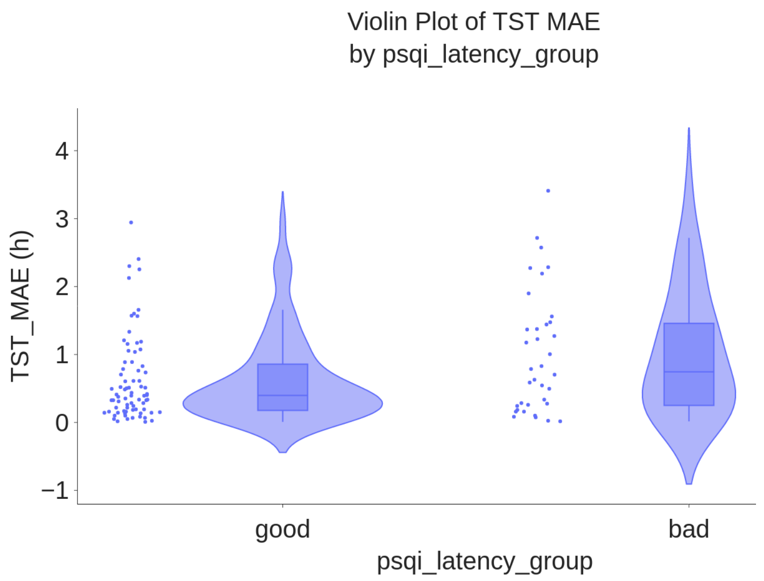


*Figure D.16: Boxplot of TST MAE (h) by PSQI sleep latency group on 117 subjects*

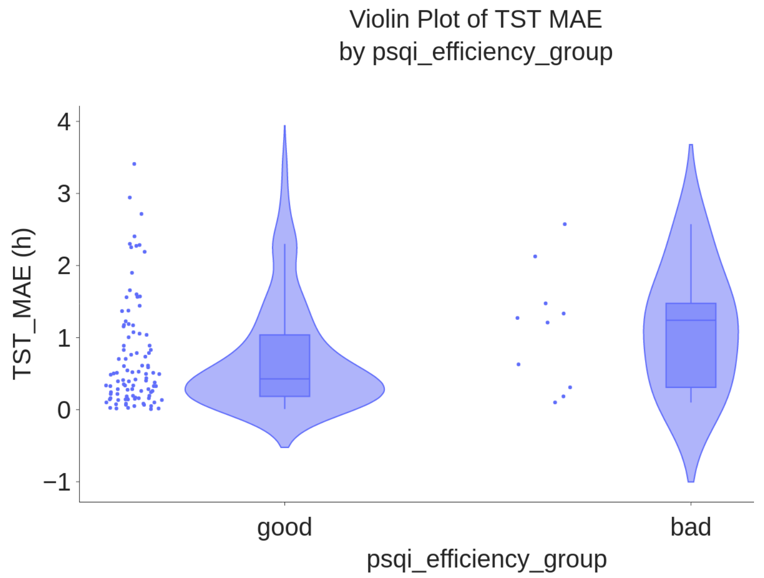

*Figure D.17: Boxplot of TST MAE (h) by PSQI sleep efficiency group on 117 subjects*

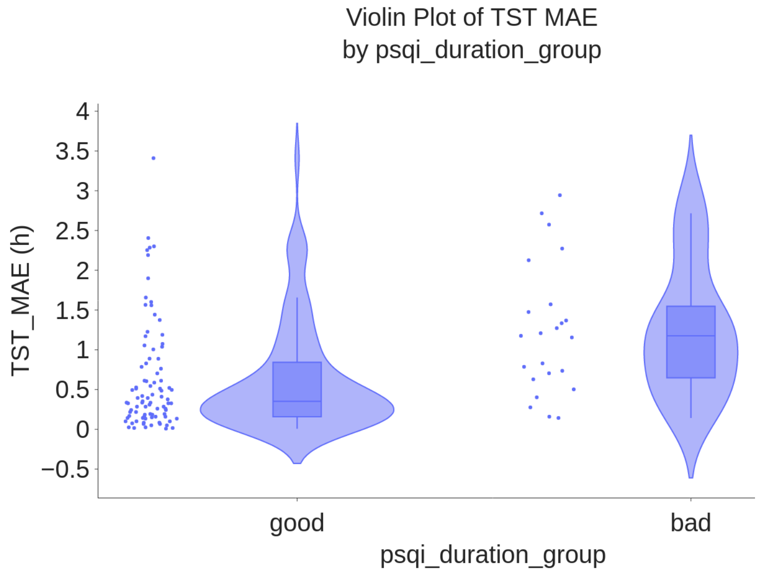


*Figure D.18: Boxplot of TST MAE (h) by PSQI sleep suration group on 117 subjects*

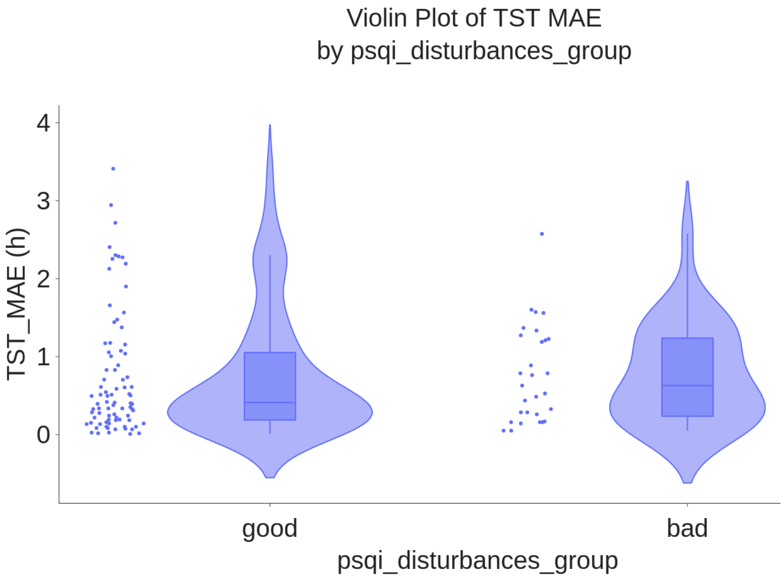

*Figure D.19: Boxplot of TST MAE (h) by PSQI sleep disturbances group on 117 subjects*

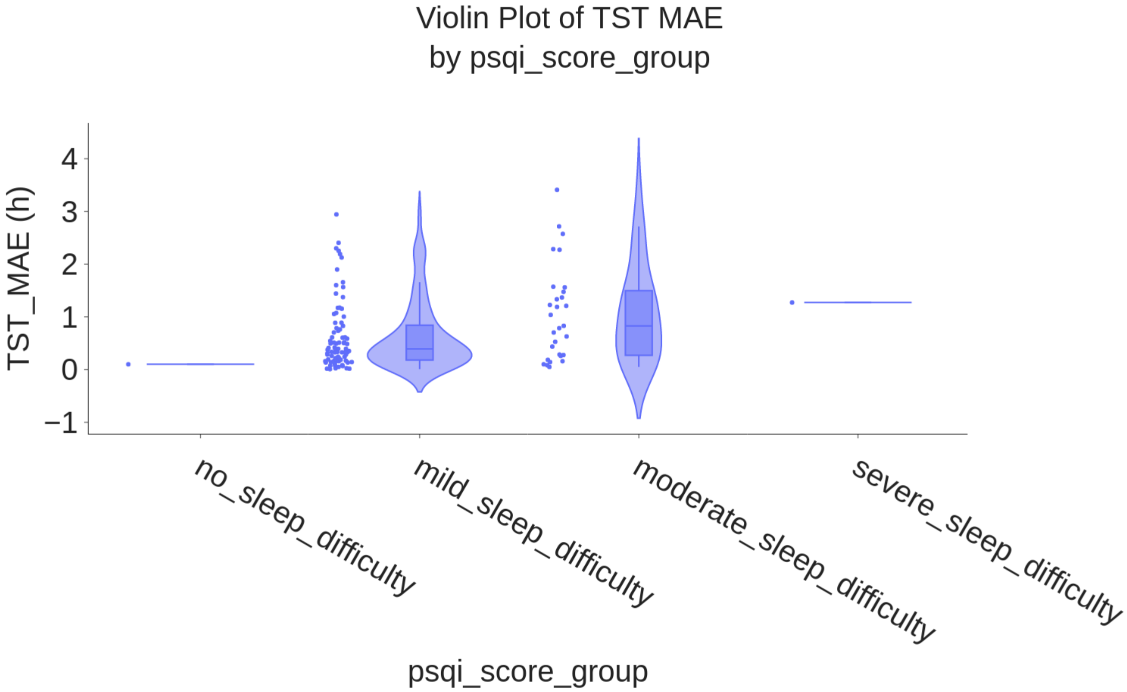


*Figure D.30: Boxplot of TST MAE (h) by PSQI score group on 117 subjects*

| Metadata | Subgroup | Number of subjects |
|---|---|---|
| bmi_group | <18.5 | 2 |
| mattress | unknown | 2 |
| hypnotics | yes | 1 |
| psqi_score_group | no_sleep_difficulty | 1 |
| psqi_score_group | severy_sleep_difficulty | 1 |
| sleep_quality | very_good | 1 |

*Table D.1: Subgroup Distribution of Participants with Insufficient Sample Sizes for Statistical Analysis*
