## Supplementary Materials E for "Evaluation of a Contactless Sleep Monitoring Device for Sleep Stage Detection against Home Polysomnography in a Healthy Population"

This table contains the results of statistical tests Kruskal-Wallis and Brown-Forsythe tests followed by Holm-Bonferroni correction.

**Legend**:
metrics (column A): Acronym of the metric being tested.
metadata (column B): Metadata category being analyzed for subgroup analysis.
alpha (column C): Significance level used for the tests, set at 0.05.
k_stat (column D): Kruskal-Wallis test statistic.
k_p_value (column E): P-value associated with the Kruskal-Wallis test statistic.
b_stat (column F): Brown-Forsythe test statistic.
b_p_value (column G): P-value associated with the Brown-Forsythe test statistic..
adjusted_k_alpha (column H): Adjusted alpha levels for the Kruskal-Wallis tests after applying the Holm-Bonferroni correction.
k_rejected (column I): Boolean indicating whether the null hypothesis was rejected for each Kruskal-Wallis test after correction. True means the null hypothesis was rejected, indicating a significant difference.
b_p_values (column J): List of p-values from the Brown-Forsythe tests before any correction for multiple comparisons.
adjusted_b_alpha (column K): Adjusted alpha levels for the Brown-Forsythe tests after applying the Holm-Bonferroni correction.
b_rejected (column L): Boolean list indicating whether the null hypothesis was rejected for each Brown-Forsythe test after correction. True means the null hypothesis was rejected, indicating a significant difference in variances.

| metrics | metadata | alpha | k_stat | k_p_value | b_stat | b_p_value | adjusted_k_alpha | k_rejected | b_p_values | adjusted_b_alpha | b_rejected |
|---|---|---|---|---|---|---|---|---|---|---|---|
| **TST** | bmi_group | 0.05 | 3.7E+00 | 3.0E-01 | 5.3E-01 | 6.7E-01 | 3.3E-04 | FALSE | 6.7E-01 | 6.8E-04 | FALSE |
| **TST** | sex | 0.05 | 3.8E-01 | 5.4E-01 | 1.0E+00 | 3.1E-01 | 5.8E-04 | FALSE | 3.1E-01 | 2.9E-04 | FALSE |
| **TST** | psqi_score_group | 0.05 | 8.0E+00 | 4.6E-02 | 2.2E+00 | 9.0E-02 | 2.3E-04 | FALSE | 9.0E-02 | 2.3E-04 | FALSE |
| **TST** | psqi_quality_group | 0.05 | 4.5E+00 | 3.4E-02 | 3.7E+00 | 5.7E-02 | 2.2E-04 | FALSE | 5.7E-02 | 2.2E-04 | FALSE |
| **TST** | psqi_latency_group | 0.05 | 3.9E+00 | 4.9E-02 | 6.6E+00 | 1.1E-02 | 2.3E-04 | FALSE | 1.1E-02 | 2.1E-04 | FALSE |
| **TST** | psqi_duration_group | 0.05 | 1.4E+01 | 1.9E-04 | 2.0E+00 | 1.6E-01 | 2.1E-04 | **TRUE** | 1.6E-01 | 2.5E-04 | FALSE |
| **TST** | psqi_efficiency_group | 0.05 | 2.7E+00 | 9.8E-02 | 4.2E-01 | 5.2E-01 | 2.6E-04 | FALSE | 5.2E-01 | 4.6E-04 | FALSE |
| **TST** | psqi_disturbances_group | 0.05 | 7.4E-01 | 3.9E-01 | 8.9E-02 | 7.7E-01 | 4.2E-04 | FALSE | 7.7E-01 | 1.0E-03 | FALSE |
| **TST** | psqi_medication_group | 0.05 | 1.6E-01 | 6.9E-01 | 2.3E-01 | 6.3E-01 | 9.6E-04 | FALSE | 6.3E-01 | 6.3E-04 | FALSE |
| **TST** | psqi_day_dysfunction_group | 0.05 | 5.2E+00 | 2.3E-02 | 1.8E-01 | 6.7E-01 | 2.2E-04 | FALSE | 6.7E-01 | 7.0E-04 | FALSE |
| **TST** | mattress | 0.05 | 1.5E+00 | 9.1E-01 | 1.1E+00 | 3.9E-01 | 3.6E-03 | FALSE | 3.9E-01 | 3.4E-04 | FALSE |
| **TST** | mattress_thickness_group | 0.05 | 3.1E+00 | 7.6E-02 | 6.9E-03 | 9.3E-01 | 2.4E-04 | FALSE | 9.3E-01 | 3.6E-03 | FALSE |
| **TST** | is_couple | 0.05 | 1.1E-03 | 9.7E-01 | 2.1E-03 | 9.6E-01 | 8.3E-03 | FALSE | 9.6E-01 | 7.1E-03 | FALSE |
| **TST** | age_group | 0.05 | 5.6E+00 | 6.0E-02 | 8.7E-01 | 4.2E-01 | 2.4E-04 | FALSE | 4.2E-01 | 3.6E-04 | FALSE |
| **TILight** | bmi_group | 0.05 | 9.2E+00 | 2.7E-02 | 7.2E-01 | 5.4E-01 | 2.2E-04 | FALSE | 5.4E-01 | 4.7E-04 | FALSE |
| **TILight** | sex | 0.05 | 3.1E+00 | 7.8E-02 | 3.4E-02 | 8.5E-01 | 2.4E-04 | FALSE | 8.5E-01 | 1.8E-03 | FALSE |
| **TILight** | psqi_score_group | 0.05 | 2.7E+00 | 4.3E-01 | 1.1E+00 | 3.6E-01 | 4.7E-04 | FALSE | 3.6E-01 | 3.3E-04 | FALSE |
| **TILight** | psqi_quality_group | 0.05 | 1.4E+00 | 2.4E-01 | 2.2E-04 | 9.9E-01 | 3.0E-04 | FALSE | 9.9E-01 | 1.7E-02 | FALSE |
| **TILight** | psqi_latency_group | 0.05 | 5.6E-02 | 8.1E-01 | 1.5E-02 | 9.0E-01 | 1.7E-03 | FALSE | 9.0E-01 | 2.6E-03 | FALSE |
| **TILight** | psqi_duration_group | 0.05 | 2.2E-01 | 6.4E-01 | 4.7E-01 | 5.0E-01 | 8.1E-04 | FALSE | 5.0E-01 | 4.2E-04 | FALSE |
| **TILight** | psqi_efficiency_group | 0.05 | 2.0E+00 | 1.6E-01 | 8.4E-01 | 3.6E-01 | 2.7E-04 | FALSE | 3.6E-01 | 3.2E-04 | FALSE |
| **TILight** | psqi_disturbances_group | 0.05 | 3.5E+00 | 6.0E-02 | 5.4E-01 | 4.6E-01 | 2.4E-04 | FALSE | 4.6E-01 | 4.1E-04 | FALSE |
| **TILight** | psqi_medication_group | 0.05 | 1.1E+00 | 3.0E-01 | 6.7E-01 | 4.1E-01 | 3.4E-04 | FALSE | 4.1E-01 | 3.6E-04 | FALSE |
| **TILight** | psqi_day_dysfunction_group | 0.05 | 4.0E-02 | 8.4E-01 | 2.0E-01 | 6.6E-01 | 1.9E-03 | FALSE | 6.6E-01 | 6.7E-04 | FALSE |
| **TILight** | mattress | 0.05 | 2.0E+01 | 1.4E-03 | 1.1E+00 | 3.8E-01 | 2.1E-04 | FALSE | 3.8E-01 | 3.4E-04 | FALSE |
| **TILight** | mattress_thickness_group | 0.05 | 1.3E-01 | 7.2E-01 | 4.8E+00 | 3.0E-02 | 1.1E-03 | FALSE | 3.0E-02 | 2.1E-04 | FALSE |
| **TILight** | is_couple | 0.05 | 7.9E-01 | 3.7E-01 | 6.3E-01 | 4.3E-01 | 4.0E-04 | FALSE | 4.3E-01 | 3.7E-04 | FALSE |
| **TILight** | age_group | 0.05 | 2.1E+00 | 3.6E-01 | 6.6E-01 | 5.2E-01 | 3.8E-04 | FALSE | 5.2E-01 | 4.5E-04 | FALSE |
| **TIDeep** | bmi_group | 0.05 | 2.2E+00 | 5.3E-01 | 1.3E+00 | 2.7E-01 | 5.6E-04 | FALSE | 2.7E-01 | 2.8E-04 | FALSE |
| **TIDeep** | sex | 0.05 | 3.6E-01 | 5.5E-01 | 3.7E+00 | 5.6E-02 | 6.0E-04 | FALSE | 5.6E-02 | 2.2E-04 | FALSE |
| **TIDeep** | psqi_score_group | 0.05 | 2.9E+00 | 4.1E-01 | 1.2E+00 | 3.0E-01 | 4.5E-04 | FALSE | 3.0E-01 | 2.9E-04 | FALSE |
| **TIDeep** | psqi_quality_group | 0.05 | 1.7E-01 | 6.8E-01 | 9.7E-02 | 7.6E-01 | 9.1E-04 | FALSE | 7.6E-01 | 9.6E-04 | FALSE |
| **TIDeep** | psqi_latency_group | 0.05 | 7.2E-02 | 7.9E-01 | 2.4E-04 | 9.9E-01 | 1.3E-03 | FALSE | 9.9E-01 | 1.3E-02 | FALSE |
| **TIDeep** | psqi_duration_group | 0.05 | 5.8E-02 | 8.1E-01 | 1.6E-01 | 6.9E-01 | 1.6E-03 | FALSE | 6.9E-01 | 7.5E-04 | FALSE |
| **TIDeep** | psqi_efficiency_group | 0.05 | 2.4E-01 | 6.2E-01 | 2.2E-01 | 6.4E-01 | 7.1E-04 | FALSE | 6.4E-01 | 6.3E-04 | FALSE |
| **TIDeep** | psqi_disturbances_group | 0.05 | 2.4E+00 | 1.2E-01 | 6.0E-01 | 4.4E-01 | 2.7E-04 | FALSE | 4.4E-01 | 3.8E-04 | FALSE |
| **TIDeep** | psqi_medication_group | 0.05 | 2.0E+00 | 1.6E-01 | 8.4E-01 | 3.6E-01 | 2.8E-04 | FALSE | 3.6E-01 | 3.3E-04 | FALSE |
| **TIDeep** | psqi_day_dysfunction_group | 0.05 | 2.7E-03 | 9.6E-01 | 3.1E-01 | 5.8E-01 | 6.3E-03 | FALSE | 5.8E-01 | 5.2E-04 | FALSE |
| **TIDeep** | mattress | 0.05 | 1.4E+01 | 1.8E-02 | 2.4E+00 | 4.4E-02 | 2.2E-04 | FALSE | 4.4E-02 | 2.2E-04 | FALSE |
| **TIDeep** | mattress_thickness_group | 0.05 | 1.5E+00 | 2.2E-01 | 6.8E-02 | 8.0E-01 | 3.0E-04 | FALSE | 8.0E-01 | 1.1E-03 | FALSE |
| **TIDeep** | is_couple | 0.05 | 1.6E+00 | 2.0E-01 | 1.4E+00 | 2.5E-01 | 2.9E-04 | FALSE | 2.5E-01 | 2.6E-04 | FALSE |
| **TIDeep** | age_group | 0.05 | 7.2E-01 | 7.0E-01 | 1.1E+00 | 3.4E-01 | 9.8E-04 | FALSE | 3.4E-01 | 3.2E-04 | FALSE |
| **TIREM** | bmi_group | 0.05 | 5.2E+00 | 1.6E-01 | 8.9E-01 | 4.5E-01 | 2.7E-04 | FALSE | 4.5E-01 | 3.9E-04 | FALSE |
| **TIREM** | sex | 0.05 | 7.7E-04 | 9.8E-01 | 2.0E-01 | 6.6E-01 | 1.0E-02 | FALSE | 6.6E-01 | 6.8E-04 | FALSE |
| **TIREM** | psqi_score_group | 0.05 | 1.7E+00 | 6.3E-01 | 1.1E+00 | 3.7E-01 | 7.4E-04 | FALSE | 3.7E-01 | 3.4E-04 | FALSE |
| **TIREM** | psqi_quality_group | 0.05 | 3.6E-01 | 5.5E-01 | 3.1E-04 | 9.9E-01 | 6.1E-04 | FALSE | 9.9E-01 | 1.0E-02 | FALSE |
| **TIREM** | psqi_latency_group | 0.05 | 1.3E+00 | 2.5E-01 | 2.5E-01 | 6.2E-01 | 3.1E-04 | FALSE | 6.2E-01 | 6.0E-04 | FALSE |
| **TIREM** | psqi_duration_group | 0.05 | 2.9E+00 | 8.7E-02 | 7.1E-05 | 9.9E-01 | 2.5E-04 | FALSE | 9.9E-01 | 2.5E-02 | FALSE |
| **TIREM** | psqi_efficiency_group | 0.05 | 2.9E+00 | 8.8E-02 | 2.3E+00 | 1.3E-01 | 2.5E-04 | FALSE | 1.3E-01 | 2.4E-04 | FALSE |
| **TIREM** | psqi_disturbances_group | 0.05 | 1.7E-02 | 9.0E-01 | 2.5E-01 | 6.2E-01 | 3.1E-03 | FALSE | 6.2E-01 | 6.0E-04 | FALSE |
| **TIREM** | psqi_medication_group | 0.05 | 4.6E-01 | 5.0E-01 | 2.8E-01 | 6.0E-01 | 5.3E-04 | FALSE | 6.0E-01 | 5.6E-04 | FALSE |
| **TIREM** | psqi_day_dysfunction_group | 0.05 | 2.2E-01 | 6.4E-01 | 3.0E-01 | 5.9E-01 | 7.9E-04 | FALSE | 5.9E-01 | 5.4E-04 | FALSE |
| **TIREM** | mattress | 0.05 | 1.1E+00 | 9.6E-01 | 3.4E-01 | 8.9E-01 | 5.6E-03 | FALSE | 8.9E-01 | 2.3E-03 | FALSE |
| **TIREM** | mattress_thickness_group | 0.05 | 4.5E+00 | 3.4E-02 | 1.5E+00 | 2.2E-01 | 2.2E-04 | FALSE | 2.2E-01 | 2.6E-04 | FALSE |
| **TIREM** | is_couple | 0.05 | 1.2E-04 | 9.9E-01 | 1.5E+00 | 2.2E-01 | 1.7E-02 | FALSE | 2.2E-01 | 2.6E-04 | FALSE |
| **TIREM** | age_group | 0.05 | 2.8E+00 | 2.5E-01 | 5.7E-01 | 5.7E-01 | 3.1E-04 | FALSE | 5.7E-01 | 5.1E-04 | FALSE |
| **PILight** | bmi_group | 0.05 | 6.7E+00 | 8.1E-02 | 2.8E-01 | 8.4E-01 | 2.4E-04 | FALSE | 8.4E-01 | 1.5E-03 | FALSE |
| **PILight** | sex | 0.05 | 7.2E-01 | 4.0E-01 | 8.2E-01 | 3.7E-01 | 4.3E-04 | FALSE | 3.7E-01 | 3.4E-04 | FALSE |
| **PILight** | psqi_score_group | 0.05 | 3.2E+00 | 3.6E-01 | 1.2E+00 | 3.2E-01 | 3.8E-04 | FALSE | 3.2E-01 | 3.0E-04 | FALSE |
| **PILight** | psqi_quality_group | 0.05 | 6.7E-01 | 4.1E-01 | 3.8E-02 | 8.5E-01 | 4.5E-04 | FALSE | 8.5E-01 | 1.5E-03 | FALSE |
| **PILight** | psqi_latency_group | 0.05 | 1.4E-02 | 9.1E-01 | 8.5E-01 | 3.6E-01 | 3.3E-03 | FALSE | 3.6E-01 | 3.2E-04 | FALSE |
| **PILight** | psqi_duration_group | 0.05 | 2.0E+00 | 1.6E-01 | 5.2E-02 | 8.2E-01 | 2.8E-04 | FALSE | 8.2E-01 | 1.3E-03 | FALSE |
| **PILight** | psqi_efficiency_group | 0.05 | 7.9E-01 | 3.7E-01 | 2.5E+00 | 1.2E-01 | 4.0E-04 | FALSE | 1.2E-01 | 2.4E-04 | FALSE |
| **PILight** | psqi_disturbances_group | 0.05 | 1.3E+00 | 2.6E-01 | 1.4E-03 | 9.7E-01 | 3.1E-04 | FALSE | 9.7E-01 | 8.3E-03 | FALSE |
| **PILight** | psqi_medication_group | 0.05 | 1.2E+00 | 2.7E-01 | 1.9E-01 | 6.7E-01 | 3.2E-04 | FALSE | 6.7E-01 | 6.9E-04 | FALSE |
| **PILight** | psqi_day_dysfunction_group | 0.05 | 5.2E-02 | 8.2E-01 | 1.7E-01 | 6.8E-01 | 1.7E-03 | FALSE | 6.8E-01 | 7.4E-04 | FALSE |
| **PILight** | mattress | 0.05 | 2.1E+01 | 7.2E-04 | 8.1E-01 | 5.4E-01 | 2.1E-04 | FALSE | 5.4E-01 | 4.8E-04 | FALSE |
| **PILight** | mattress_thickness_group | 0.05 | 1.0E+00 | 3.1E-01 | 1.9E+00 | 1.7E-01 | 3.4E-04 | FALSE | 1.7E-01 | 2.5E-04 | FALSE |
| **PILight** | is_couple | 0.05 | 2.3E+00 | 1.3E-01 | 8.4E-02 | 7.7E-01 | 2.7E-04 | FALSE | 7.7E-01 | 1.0E-03 | FALSE |
| **PILight** | age_group | 0.05 | 5.1E+00 | 8.0E-02 | 2.8E-01 | 7.6E-01 | 2.4E-04 | FALSE | 7.6E-01 | 9.8E-04 | FALSE |
| **PIDeep** | bmi_group | 0.05 | 1.5E+00 | 6.9E-01 | 9.3E-01 | 4.3E-01 | 9.4E-04 | FALSE | 4.3E-01 | 3.7E-04 | FALSE |
| **PIDeep** | sex | 0.05 | 8.3E-01 | 3.6E-01 | 3.7E+00 | 5.7E-02 | 3.8E-04 | FALSE | 5.7E-02 | 2.2E-04 | FALSE |
| **PIDeep** | psqi_score_group | 0.05 | 3.1E+00 | 3.8E-01 | 1.6E+00 | 1.9E-01 | 4.1E-04 | FALSE | 1.9E-01 | 2.5E-04 | FALSE |
| **PIDeep** | psqi_quality_group | 0.05 | 2.4E-01 | 6.3E-01 | 2.3E-01 | 6.3E-01 | 7.2E-04 | FALSE | 6.3E-01 | 6.2E-04 | FALSE |
| **PIDeep** | psqi_latency_group | 0.05 | 1.3E-01 | 7.2E-01 | 7.9E-03 | 9.3E-01 | 1.1E-03 | FALSE | 9.3E-01 | 3.3E-03 | FALSE |
| **PIDeep** | psqi_duration_group | 0.05 | 1.8E-01 | 6.7E-01 | 2.4E-03 | 9.6E-01 | 8.9E-04 | FALSE | 9.6E-01 | 6.3E-03 | FALSE |
| **PIDeep** | psqi_efficiency_group | 0.05 | 2.0E-01 | 6.5E-01 | 1.8E+00 | 1.8E-01 | 8.5E-04 | FALSE | 1.8E-01 | 2.5E-04 | FALSE |
| **PIDeep** | psqi_disturbances_group | 0.05 | 5.3E+00 | 2.2E-02 | 2.9E+00 | 9.1E-02 | 2.2E-04 | FALSE | 9.1E-02 | 2.3E-04 | FALSE |
| **PIDeep** | psqi_medication_group | 0.05 | 1.7E+00 | 1.9E-01 | 2.1E+00 | 1.5E-01 | 2.9E-04 | FALSE | 1.5E-01 | 2.4E-04 | FALSE |
| **PIDeep** | psqi_day_dysfunction_group | 0.05 | 8.9E-03 | 9.2E-01 | 1.9E+00 | 1.7E-01 | 4.2E-03 | FALSE | 1.7E-01 | 2.5E-04 | FALSE |
| **PIDeep** | mattress | 0.05 | 2.5E+01 | 1.3E-04 | 2.0E+00 | 8.2E-02 | 2.1E-04 | **TRUE** | 8.2E-02 | 2.3E-04 | FALSE |
| **PIDeep** | mattress_thickness_group | 0.05 | 7.8E-01 | 3.8E-01 | 3.3E-01 | 5.7E-01 | 4.1E-04 | FALSE | 5.7E-01 | 5.0E-04 | FALSE |
| **PIDeep** | is_couple | 0.05 | 7.6E-01 | 3.8E-01 | 9.2E-01 | 3.4E-01 | 4.2E-04 | FALSE | 3.4E-01 | 3.1E-04 | FALSE |
| **PIDeep** | age_group | 0.05 | 7.5E-01 | 6.9E-01 | 8.3E-01 | 4.4E-01 | 9.3E-04 | FALSE | 4.4E-01 | 3.8E-04 | FALSE |
| **PIREM** | bmi_group | 0.05 | 3.5E+00 | 3.2E-01 | 1.3E+00 | 2.7E-01 | 3.6E-04 | FALSE | 2.7E-01 | 2.7E-04 | FALSE |
| **PIREM** | sex | 0.05 | 2.9E-01 | 5.9E-01 | 1.1E-01 | 7.4E-01 | 6.4E-04 | FALSE | 7.4E-01 | 8.6E-04 | FALSE |
| **PIREM** | psqi_score_group | 0.05 | 3.2E+00 | 3.6E-01 | 9.3E-01 | 4.3E-01 | 3.9E-04 | FALSE | 4.3E-01 | 3.7E-04 | FALSE |
| **PIREM** | psqi_quality_group | 0.05 | 1.4E+00 | 2.4E-01 | 9.8E-02 | 7.6E-01 | 3.0E-04 | FALSE | 7.6E-01 | 9.4E-04 | FALSE |
| **PIREM** | psqi_latency_group | 0.05 | 2.9E+00 | 8.8E-02 | 3.7E-02 | 8.5E-01 | 2.5E-04 | FALSE | 8.5E-01 | 1.7E-03 | FALSE |
| **PIREM** | psqi_duration_group | 0.05 | 3.8E+00 | 5.2E-02 | 2.8E-03 | 9.6E-01 | 2.3E-04 | FALSE | 9.6E-01 | 5.6E-03 | FALSE |
| **PIREM** | psqi_efficiency_group | 0.05 | 3.5E+00 | 6.0E-02 | 3.0E-01 | 5.9E-01 | 2.4E-04 | FALSE | 5.9E-01 | 5.4E-04 | FALSE |
| **PIREM** | psqi_disturbances_group | 0.05 | 4.2E-01 | 5.2E-01 | 1.1E+00 | 2.9E-01 | 5.4E-04 | FALSE | 2.9E-01 | 2.8E-04 | FALSE |
| **PIREM** | psqi_medication_group | 0.05 | 6.2E-02 | 8.0E-01 | 9.7E-01 | 3.3E-01 | 1.5E-03 | FALSE | 3.3E-01 | 3.0E-04 | FALSE |
| **PIREM** | psqi_day_dysfunction_group | 0.05 | 8.9E-03 | 9.2E-01 | 1.4E-01 | 7.0E-01 | 4.5E-03 | FALSE | 7.0E-01 | 7.8E-04 | FALSE |
| **PIREM** | mattress | 0.05 | 2.3E+00 | 8.0E-01 | 3.7E-01 | 8.7E-01 | 1.5E-03 | FALSE | 8.7E-01 | 2.0E-03 | FALSE |
| **PIREM** | mattress_thickness_group | 0.05 | 2.8E+00 | 9.7E-02 | 2.1E+00 | 1.5E-01 | 2.6E-04 | FALSE | 1.5E-01 | 2.4E-04 | FALSE |
| **PIREM** | is_couple | 0.05 | 6.8E-01 | 4.1E-01 | 4.9E-05 | 9.9E-01 | 4.4E-04 | FALSE | 9.9E-01 | 5.0E-02 | FALSE |
| **PIREM** | age_group | 0.05 | 4.5E-01 | 8.0E-01 | 3.1E-01 | 7.4E-01 | 1.4E-03 | FALSE | 7.4E-01 | 8.8E-04 | FALSE |
