## Supplementary Materials F for "Evaluation of a Contactless Sleep Monitoring Device for Sleep Stage Detection against Home Polysomnography in a Healthy Population"

| **Legend**<br>EBE: Epoch by epoch<br>S-W: Sleep-Wake<br><br>**Results must be interpreted with cautious as the dataset used for WSA performance evaluation has been collected in real-life conditions, is larger and more diverse than the other datasets according to the protocol described in other studies.**<br><br>**For all these reasons, the reader must keep in mind that the evaluation of the other devices on the associated datasets may overestimate their performance compared to the results obtained for WSA we provide in this study.**<br><br>**The performance presented in this document of the listed devices has been computed based on the confusion matrix when it was available, since the data for each night were not provided. This explains the discrepancies between the performance of Withings Sleep Analyzer in this document and in the full-text.** | | **General Information** | | | | | | **EBE metrics: S-W** | | | | | | **EBE metrics: sleep stages** | | | | |
|---|---|---|---|---|---|---|---|---|---|---|---|---|---|---|---|---|---|---|
| **Reference** | **Device** | **Wearable/Nearable** | **# subjects** | **Reference** | **Study setting** | **# nights** | **Comment** | **Mean reference TST (in minutes) ± std** | **Se sleep / Sp wake** | **Sp sleep / Se wake** | **Accuracy** | **Proportion of wake data (%)** | **Kappa** | **Se light** | **Se deep** | **Se REM** | **Accuracy** | **Kappa** |
| **Chinoy2021** | Philips Respironics Actiwatch 2 | W | 34 healthy young adults | PSG<br>(Siesta; Compumedics) | Sleep Lab<br>3 nights per subject | 98 | **Exclusions post collection**<br>We selected a duration of <470 min (ie, <7h30 min) a | 418 ± 41 | 0.91 | 0.63 | 0.89 | 8 | 0.42 | N/A | N/A | N/A | N/A | N/A |
| | Fatigue Science Readiband | W | 15 healthy young adults | | | 41 | | 416 ± 48 | 0.92 | 0.55 | 0.88 | 13 | 0.43 | | | | | |
| | Fitbit Alta HR | W | 20 healthy young adults | | | 49 | | 425 ± 33 | 0.95 | 0.54 | 0.90 | 11 | 0.51 | 0.76 | 0.53 | 0.69 | 0.68 | 0.51 |
| | Garmin Fenix 5S | W | 11 healthy young adults | | | 29 | | 413 ± 53 | 0.99 | 0.18 | 0.88 | 14 | 0.25 | 0.68 | 0.56 | 0.50 | 0.55 | 0.30 |
| | Garmin Vivosmart 3 | W | 15 healthy young adults | | | 43 | | 415 ± 48 | 0.99 | 0.19 | 0.88 | 14 | 0.26 | 0.70 | 0.56 | 0.54 | 0.58 | 0.33 |
| | EarlySense Live | N | 19 healthy young adults | | | 51 | | 422 ± 35 | 0.96 | 0.47 | 0.90 | 12 | 0.48 | 0.57 | 0.68 | 0.64 | 0.59 | 0.40 |
| | ResMed S+ | N | 19 healthy young adults | | | 51 | | 422 ± 34 | 0.93 | 0.51 | 0.88 | 12 | 0.44 | 0.67 | 0.59 | 0.50 | 0.60 | 0.39 |
| | SleepScore Max | N | 15 healthy young adults | | | 42 | | 414 ± 49 | 0.94 | 0.50 | 0.88 | 14 | 0.46 | 0.68 | 0.59 | 0.49 | 0.60 | 0.39 |
| **Kainec2024** | Philips Repironics Actiwatch Spectrum Plus | W | 53 healthy young adults | PSG<br>(EasyCap for 33 participants<br>&<br>EasyCap + BrainAmp MR plus amplifiers, Brain Products GmbH for 20 other participants) | Sleep Lab | 53 | **Device position**<br>The Fitbit Versa and Garmin Vivosmart were applied<br><br>**Short nights exclusion**<br>Three participants slept less than 4.5 h overnight in th<br><br>**Heteroscedasticity**<br>Briefly, plots were examined for heteroscedasticity an | 435 ± 57 | N/A | N/A | N/A | N/A | N/A | N/A | N/A | N/A | N/A | N/A |
| | Fitbit Inspire HR | W | | | | 53 | | 434 ± 53 | | | | | | | | | | |
| | Fitbit Versa 2 | W | | | | 53 | | 435 ± 57 | | | | | | | | | | |
| | Garmin Vivosmart 4 | W | | | | 53 | | 435 ± 54 | | | | | | | | | | |
| | Oura Ring | W | | | | 53 | | 435 ± 54 | | | | | | | | | | |
| | Withings Sleep Analyzer | N | | | | 53 | | 436 ± 56 | | | | | | | | | | |
| **Miller2022** | Apple Watch S6 | W | 53 healthy young adults | PSG<br>Grass gold-cup electrodes<br>(AstroMed) | Sleep Lab | 1/subject | **Start/end time**<br>For all wearable devices other than Somfit, the start a<br>onset or after each device's self-determined sleep off | N/A | 0.96 | 0.26 | 0.73 | 33 | 0.27 | 0.44 | 0.71 | | 0.47 | 0.20 |
| | Garmin Forerunner 245 | W | | | | | | | 0.98 | 0.27 | 0.81 | 25 | 0.33 | 0.68 | 0.28 | 0.50 | 0.43 | 0.24 |
| | Polar Vantage V | W | | | | | | | 0.93 | 0.51 | 0.82 | 25 | 0.49 | 0.60 | 0.33 | 0.49 | 0.48 | 0.31 |
| | Oura Ring Generation 2 | W | | | | | | | 0.94 | 0.57 | 0.85 | 25 | 0.56 | 0.66 | 0.62 | 0.52 | 0.59 | 0.46 |
| | WHOOP 3.0 | W | | | | | | | 0.92 | 0.56 | 0.83 | 25 | 0.52 | 0.58 | 0.62 | 0.66 | 0.60 | 0.57 |
| | Somfit | W | | | | | | | 0.89 | 0.57 | 0.82 | 20 | 0.45 | 0.68 | 0.68 | 0.60 | 0.64 | 0.50 |
| **Tal2017** | EarlySense | N | 85:<br>43 in setup I<br>7 in setup II<br>13 in setup III | Full PSG<br>(Respironics Alice 5 Diagnostic Sleep System)<br><br>The home setups included one of two models of portable/partial PSG: Embletta Gold or X100 (Embla Corp, Bloomfield, CO, USA). | setup I: Sleep Lab<br>ıp II: at home with no sleep part<br>ıp III: at home with a sleep part | 1/subject | **3 setups** (in lab & at home)<br>For the home setups, we recruited healthy volunteers<br>at home (setup III) | N/A | setup I: 0.90<br>setup II: 0.95<br>setup III: 0.95 | setup I: 0.83<br>setup II: 0.72<br>setup III: 0.79 | setup I: 0.89<br>setup II: 0.92<br>setup III: 0.93 | setup I: 0.19<br>setup II: 0.14<br>setup III: 0.15 | setup I: 0.66<br>setup II: 0.68<br>setup III: 0.72 | setup i: 0.63<br>setup ii: 0.64<br>setup iii: 0.68 | setup i: 0.54<br>setup ii: 0.68<br>setup iii: 0.52 | setup i: 0.40<br>setup ii: 0.63<br>setup iii: 0.64 | setup i: 0.63<br>setup ii: 0.66<br>setup iii: 0.66 | setup I: 0.42<br>setup II: 0.49<br>setup III: 0.50 |
| **Our work** | Withings Sleep Analyzer | N | 117 healthy adults | Partial PSG (CID-LXe) | at home | 1/subject | N/A | 435 (71) | 0.93 | 0.73 | 0.87 | 29 | 0.63 | 0.68 | 0.53 | 0.54 | 0.63 | 0.50 |
